# Early stopping in clinical PET studies: how to reduce expense and exposure

**DOI:** 10.1101/2020.09.13.20192856

**Authors:** Jonas Svensson, Martin Schain, Gitte M. Knudsen, Todd Ogden, Pontus Plavén-Sigray

**Affiliations:** Department of Clinical Neuroscience, Center for Psychiatry Research, Karolinska Institutet and Stockholm County Council, SE-171 76 Stockholm, Sweden; Neurobiology Research Unit, Copenhagen University Hospital, Rigshospitalet, Copenhagen, Denmark; Institute of Clinical Medicine, University of Copenhagen, Denmark; Department of Biostatistics, Mailman School of Public Health, Columbia University, New York, NY, USA; Molecular Imaging and Neuropathology Area, New York State Psychiatric Institute, New York, NY, USA

**Keywords:** early stopping, sequential testing, bayes factor, positron emission tomography, tutorial

## Abstract

Clinical positron emission tomography (PET) research is costly and entails exposing participants to radioactivity. Researchers should therefore aim to include just the number of subjects needed to fulfill the purpose of the study. In this tutorial we show how to apply *sequential Bayes Factor testing* in order to stop the recruitment of subjects in a clinical PET study as soon as enough data have been collected to make a conclusion. By using simulations, we demonstrate that it is possible to stop a study early, while keeping the number of erroneous conclusions low. We then apply sequential Bayes Factor testing to a real PET data set and show that it is possible to obtain support in favor of an effect while simultaneously reducing the sample size with 30%. Using this procedure allows researchers to reduce expense and radioactivity exposure for a range of effect sizes relevant for PET research.

## Introduction

Positron emission tomography (PET) examinations are expensive and may impose a substantial burden on research budgets. Depending on the local PET centers finances and the experimental design, it is not unusual that researchers pay 5000 Euro/USD or more for a PET-scan of a single subject. In addition to the high cost, a PET scan entails exposing individuals to radioactivity, with average doses often ranging between 0.6 to 5 mSv^1^ (Van Der Aart et al. 2012). It is hence in the interest of the PET researcher to keep the number of included research subjects to a minimum, while still performing enough PET examinations to be able to draw appropriate conclusions from the collected data.

Traditionally, the number of included subjects in a clinical PET study is determined *a priori* by a power analysis based on the null-hypothesis-significance testing (NHST) procedure^2^ (Perezgonzalez 2015). Following this, subjects are recruited and scanned until the specified sample size is reached. In the case of a patient-control comparison study, a p-value for the mean difference of an outcome in a region of interest is then calculated. This value reflects the plausibility of obtaining the observed mean difference, or a more extreme difference, given that the null hypothesis (H_0_) is true (i.e., no difference between the groups). If the p-value is above a predefined alpha threshold (usually set to 0.05), the result is interpreted as inconclusive and H_0_ is not rejected (nor may it be confirmed!). The researcher might have committed a type II error, in which case more data would likely be needed to detect a potential difference. Alternatively, H_0_ might be true, i.e., there is no population difference to detect. If the p-value is below the alpha threshold, the result is instead interpreted as there being a significant difference between groups, inferring that there is an actual difference at the population level. Although many PET-studies likely suffer from insufficient power because of limited sample sizes, it is also important to note that a difference between groups may often be detectable before reaching the sample size that was determined *a priori*. Ideally, the researchers should include just the number of individuals needed to reach an appropriate conclusion, no more, no less. When performing more scans than needed, the PET researchers are wasting money and exposing people unnecessarily to radioactivity.

One way to avoid superfluous PET scans is to intermittently check for a statistical effect while the study is still ongoing, generally termed *sequential testing* of data. In the uncorrected NHST framework, however, sequential testing does pose a problem, as it can greatly inflate the nominal type I error rate. If a new uncorrected p-value is calculated and used for making inference after the collection of each patient-control pair, the type I error rate will be above 20% for commonly seen sample sizes in PET literature (Strube 2006; Albers 2019).

Example 1

We simulated a population of patients and controls showing no difference in mean binding potential. We then sequentially drew a random patient and a random control from the population, and calculated the ensuing p-value for the mean difference in binding potential, comparing it to an significance threshold of 0.05. This was repeated until we either reached a maximum sample size of N=20 subjects/group, or the p-value fell below the threshold - in which case data collection was stopped. Repeating this procedure many times, the type I error rate was shown to increase from 5% to an average of 22%.

There exist however techniques that are appropriate to use when performing sequential testing. One possibility involves correcting the significance threshold so that the overall error rate does not surpass the alpha level when performing intermittent NHST tests (Proschan et al. 2006) (see the Discussion for a more elaborate description of this procedure). An alternative possibility is to use an entirely different metric to test scientific hypotheses: the Bayes Factor (BF) (Jeffreys 1961; Kass and Raftery 1995; Lee and Wagenmakers 2014; Schönbrodt and Wagenmakers 2018). The BF has been gaining traction in the field of biomedicine during the last decade and has two important characteristics; first, it is well suited for sequential testing of data (Rouder 2014; Schönbrodt et al. 2017), and second, it also allows for quantification of relative evidence in favour of H_0_, meaning that a PET study can be stopped when it is determined that H_0_ is supported (Dienes 2014). In this tutorial we will show how to use sequential BF tests in common PET study designs, in order to stop data collection in a study early.

### Bayes Factor - a versatile alternative for testing hypotheses

#### Support in data for competing hypotheses

Bayesian hypothesis testing using BF aims to assess how compatible the observed data (such as a patient-control difference) is with each of two competing hypotheses. These hypotheses are often specified as the null hypothesis (H_0_) and an alternative hypothesis (H_1_). The null-hypothesis typically states that the effect is exactly zero and the alternative hypothesis states that the effect is different from zero. The BF, quantifying the support of the alternative hypothesis over the null-hypothesis, is defined as the likelihood ratio of the two hypotheses:

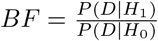

where D denotes the observed data^3^ (Wrinch and Jeffreys 1921). For example, a BF of 4 can be interpreted as “the observed data is 4 times more likely to have occurred under the alternative hypothesis compared to the null hypothesis.” As such, the BF directly quantifies the evidence in data in favour of one hypothesis against another. The reciprocal of BF, quantifies the support in data in favour of the null hypothesis, compared to the alternative. A BF of 1*/*5 would hence mean that there is 5 times more support in favor of H_0_, compared to H_1_.

#### Evidence thresholds

The BF quantifies evidence on a continuous scale ranging from 0 to infinity, where values over 1 support the hypothesis in the numerator, and values below 1 support the hypothesis in the denominator. A set of thresholds have been suggested to help with the interpretation and decision making when using BF (Table 1) (Lee and Wagenmakers 2014). A BF of 3, which often corresponds to a p-value around 0.05 (Robert and Caron 1996; Dienes 2014; Benjamin et al. 2017), is commonly interpreted as providing “moderate” evidence in favour of one hypothesis over another. It is commonly considered to be the minimal threshold for claiming support of a hypothesis^4^.

**Table 1:** Suggested evidence thresholds for the Bayes Factor, adapted from Lee & Wagenmakers, 2014.

| Bayes Factor | Interpretation |
| --- | --- |
| $>10$ | Strong evidence in favor of $H_1$ |
| $3 - 10$ | Moderate evidence in favor of $H_1$ |
| $1 - 3$ | Negligible to weak evidence in favor of $H_1$ |
| $1/3 - 1$ | Weak to negligible evidence in favor of $H_0$ |
| $1/10 - 1/3$ | Moderate evidence in favor of $H_0$ |
| $<1/10$ | Strong evidence in favor of $H_0$ |

#### Specifying the alternative hypothesis

In the classical NHST framework, the alternative hypothesis is often specified as meaning any other value than point zero. The use of BF does however require the researchers to be more specific when describing H_1_. For example, the researcher could specify the alternative hypothesis as a single value different from zero, such as predicting that a mean patient-control difference in binding potential will be exactly 0.5. However, since it is rare that researchers are confident in predicting a single point value, the alternative hypothesis is commonly specified as a probability distribution covering a range of values instead. In doing so, the researchers are “hedging their bets” by spreading the prediction out across many plausible values of an effect. This probability distribution can be informative (Stefan et al. 2019; Gronau et al. 2020), e.g., a narrow normal distribution centered around a specific value. It can also be made “non-informative,” e.g., a wide distribution centered around zero.

A commonly used non-informative distribution for describing H_1_ when testing mean differences is a two sided Cauchy distribution centered around zero^5^ (Figure 1) (Gönen et al. 2005). This distribution ranges from minus infinity to plus infinity, and has fat tails relative to other continuous distributions. Using a Cauchy distribution means that the researcher specifies H_1_ to reflect the belief that the effect (such as a patient-control difference) is of small or medium size with relatively high confidence, but also allowing, with less confidence, an effect of a larger size. The fatness of the tails is determined by the width parameter *r* (analogous to the SD of a normal distribution), and is by convention often set to 0.707. Formally, the two competing hypotheses in such a BF test are:

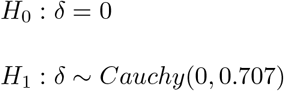

where *δ* denotes the true population effect, and “*∼*” means “distributed according to.” This particular null and alternative-hypothesis pair has become so common today when testing mean differences that it is called the “default” BF t-test (Rouder et al. 2009; Ly et al. 2016). In this article, we will only evaluate sequential testing in PET studies using the default BF t-test.

Example 2

We examined cerebral difference in [^11^C]DASB binding potential (an index of serotonin transporter availability) between patients with seasonal affective disorder and healthy control subjects in the winter (N=17 vs. 23) and summer seasons (N = 20 vs. 23) by extracting data from Figure 1 in McMahon et al., 2016. The group means were compared using a two-sided default BF t-test. In summer, there was 3 times more support in favour of H_0_ compared to H_1_ (BF = 0.32). In winter, there was instead 3 times more support in favour of H_1_ compared to H_0_ (BF = 3.01), with patients showing higher binding. We can hence conclude that there is moderate evidence of no difference in serotonin transporter availability between patients and controls in the summer season, contrary to the winter season.

**Figure 1:**
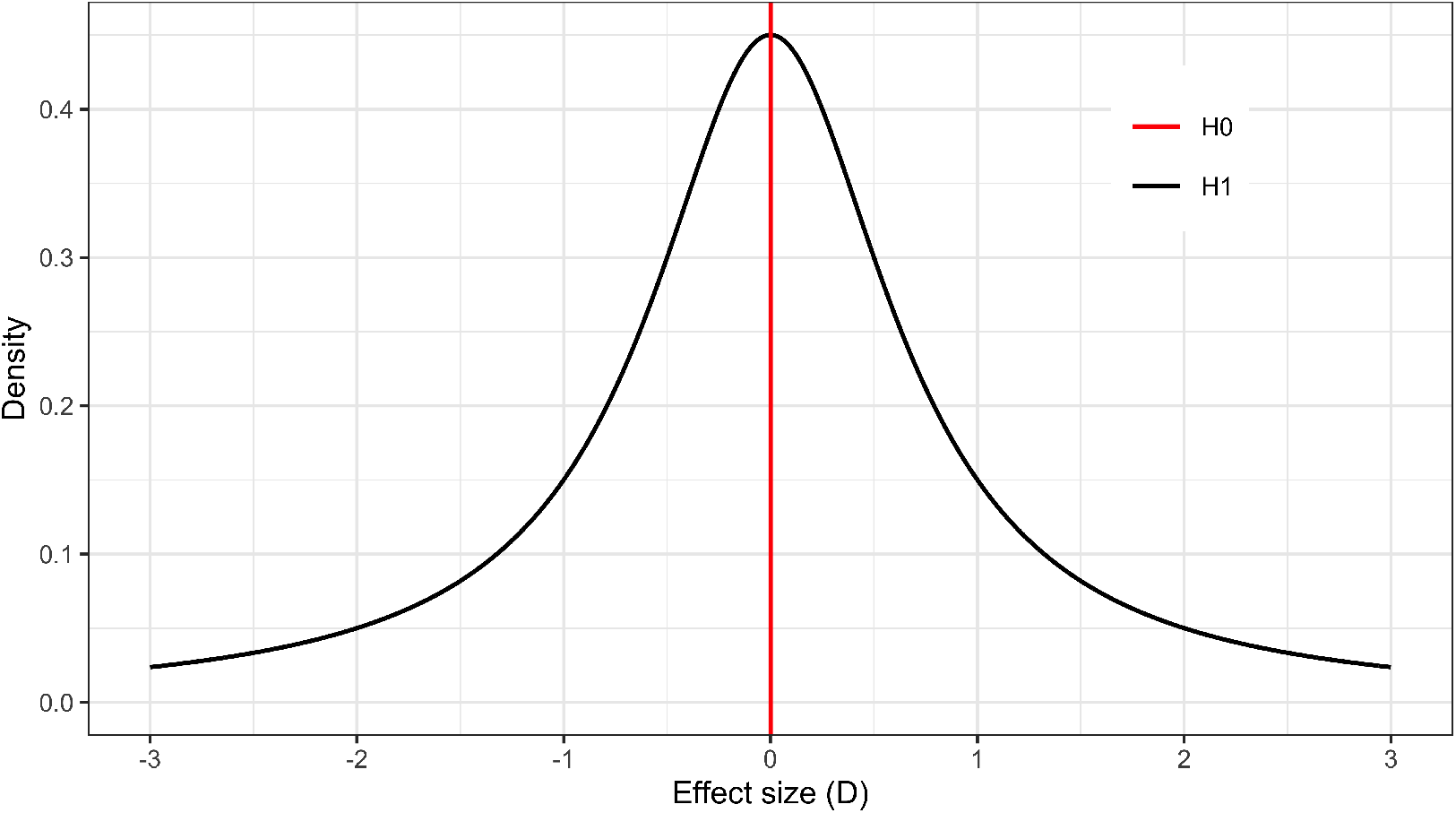
Example of two hypotheses compared in BF hypothesis testing. The alternative hypothesis (H_1_) in black is represented by a two-sided Cauchy distribution centered on zero with an r (width-parameter) of 0.707. The null hypothesis (H_0_) in red, is defined as the point zero value.

#### Sequential BF testing

In sequential BF testing, the null hypothesis is assessed in constant competition to the alternative hypothesis. If a BF is calculated after the collection of each data point, it informs the researcher how the stated hypotheses are gaining or losing support from the data. Because of this, the BF can be used for “online” monitoring of incoming data. The researchers can stop when they reach a pre-set decision threshold, a pre-set maximum sample size (or when they simply run out of money, time or patience) (Rouder 2014; Schönbrodt et al. 2017).

However, it is important to note that BF testing is subject to the same sources of uncertainty as NHST inference, i.e., the data could potentially lend support to the wrong hypothesis. This means that in sequential BF testing, the researchers can end up stopping a study early and reach an incorrect conclusion (Figure 2). In order to plan a PET study when intending to use a sequential BF design, the researchers should be aware of the different errors that can occur when making stopping decisions. False positive evidence is defined as data supporting the alternative hypothesis, when the population effect truly is zero (cf. the type I error in the classical NHST framework). False negative evidence is defined as data supporting the null hypothesis, when in fact there is a real effect in the population. It is also possible that the sequential BF testing procedure ends up unable to stop for either hypotheses, by never passing the decision threshold before a maximum possible fixed sample size (Nmax) is reached. In this tutorial, we refer to such results as “inconclusive” stopping outcomes. Figure 2 depicts the continuous evidence updating, after each added data point, and shows the three possible outcomes: a stop decision for the alternative hypothesis, a stop decision for null hypothesis and an inconclusive stop outcome, when applying sequential BF testing.

**Figure 2:**
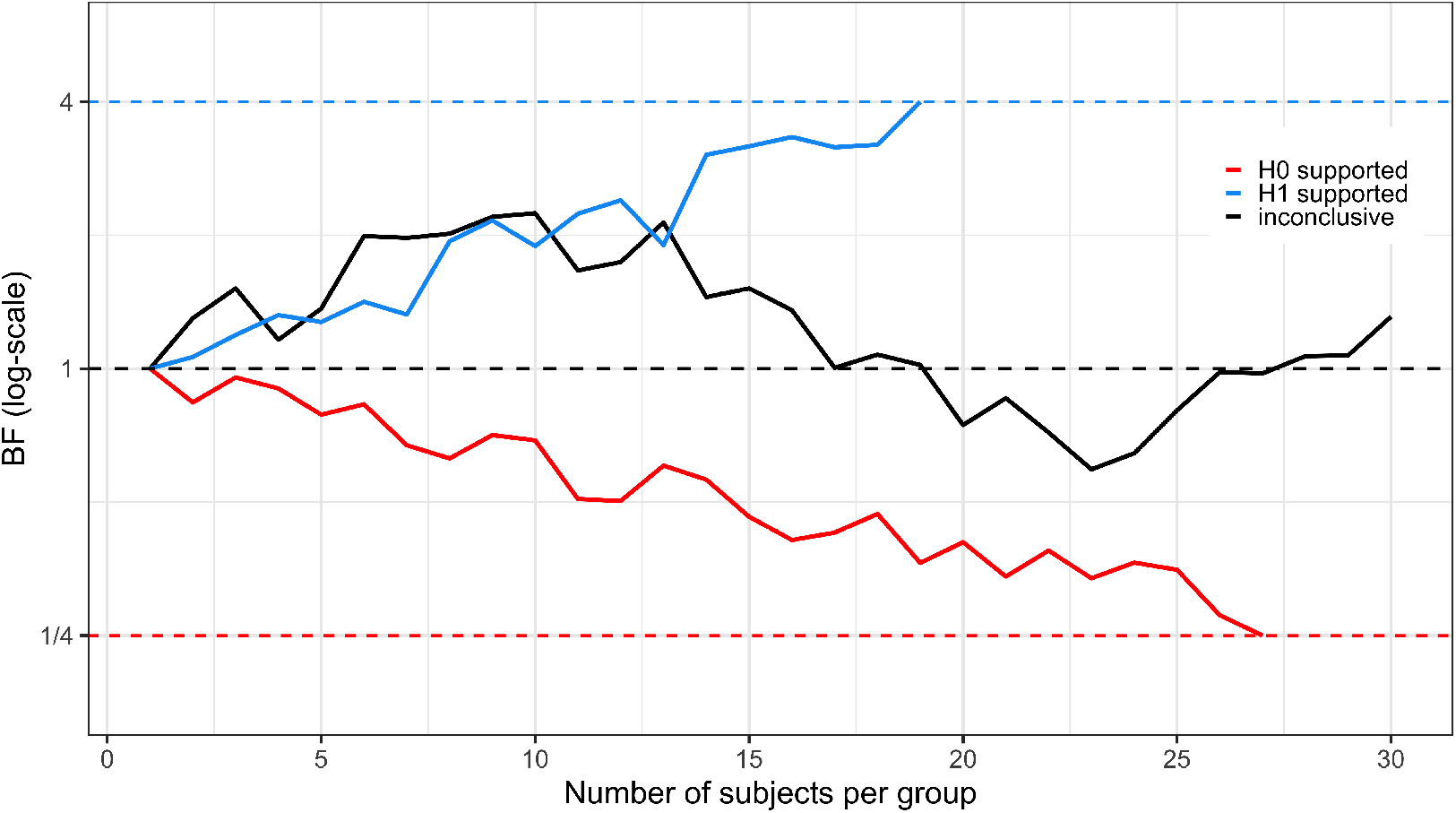
Possible outcomes of sequential testing using Bayes Factor. Three different studies have been initiated, all having N = 30 as the maximum possible sample size. For the blue line, BF crosses the pre-specified threshold (here BF > 4) triggering an early stop decision in favor of an effect. Assuming there is a true effect in the population, this would be a “true positive” finding. If there is no effect in the population, this would be a “false positive” finding. For the red line, the reciprocal of BF crosses the pre-specified threshold (here < ¼), meaning that the study is stopped in favor of the null hypothesis. Assuming there is no effect in the population this would be a “true negative”, but if there is an effect, this would be a “false negative” finding. For the black line, BF does not reach either below ¼ or above 4 before the pre-specified number of subjects have been reached, and therefore denotes an inconclusive stopping outcome

If a researcher continues to collect data indefinitely, the default BF t-test will eventually converge to supporting the hypothesis most compatible with the population effect, i.e., it will show evidence for the null-hypothesis if in fact there is no true mean difference, or for the alternative-hypothesis if there is an underlying difference in the population (Rouder et al. 2009). However, such continuous collection of data is often unrealistic in a clinical PET study. Apart from the high cost, a PET examination also entails injecting subjects with radioactivity. Hence, a maximum sample size usually has to be decided on *a priori* and approved by an ethical review board and/or a radiation safety committee.

In summary, BF is a versatile metric that can be used for different purposes. In the PET literature, there have so far only been a few articles that have used BF to e.g., complement reported p-values, as a stand-alone metric to quantify the evidence of stated hypotheses, or to assess the replicability of previously published results (Griffioen et al. 2018; Plavén-Sigray et al. 2018; Varnäs et al. 2018). Of particular interest for this tutorial, the BF first lends itself naturally to sequential testing of data (Rouder 2014; Schönbrodt et al. 2017); and second, it quantifies evidence in favour of either H_1_ and H_0_, meaning that the null-hypothesis also can gain, or lose, support by the data and hence be formally accepted or rejected (Rouder et al. 2009; Dienes 2014). However, as with all metrics used for statistical inference, the BF is not without limitations and criticism (Dienes 2008; Gelman et al. 2013).

In this tutorial we will explore how to use the default BF t-test to sequentially test data in two common clinical PET study designs: a cross-sectional (e.g., a patient-control comparison) and a paired (e.g., a pre-intervention-post scan) design. Our first aim is to demonstrate the relative merit of using sequential BF testing to stop data collection at an earlier stage, compared to simply applying a conventional “fixed N” design, while still keeping the number of false positives below the commonly set threshold of 5%. Our second aim is to assess whether sequential BF testing can be used to stop early, not only when there is an effect in the population (H_1_ is true), but also when there is no effect in the population (H_0_ is true). Our third aim is to apply sequential BF testing on real clinical PET data, and evaluate the outcome.

#### Standardized effect sizes

Throughout this tutorial we use standardized effect sizes instead of raw values of e.g., binding potential (BP_ND_), total distribution volume (V_T_) or percentage differences when we simulate mean differences. To derive a standardized effect for a comparison of two different groups, such as patients and controls, we divide the average difference in the raw outcome by the pooled standard deviation of the two groups, and call the result Cohen’s D (Cohen 1998). For a comparison within the same individual over time, such as a pre-scan-intervention-post-scan design, we divide the average difference in the raw outcome by the within-subject standard deviation (i.e., the standard deviation of the difference score), and call the result Cohen’s Dz (Lakens 2013).

The main rationale for using standardized effect sizes is that it allows us to generalize the results of the simulations to all radioligands. In Table 2 we present a subset of commonly used radioligands in PET research (Nord et al. 2014; Collste et al. 2016; Finnema et al. 2018; Svensson et al. 2019), and how differences in BP_ND_ or V_T_ translate into standardized effect sizes and vice versa.

**Table 2:** The standardized effect size used to assess the difference between two groups (Cohen’s D for a cross-sectional design) and difference within the same subjects (Cohen’s Dz for a paired design) translated into raw mean difference in BP_ND_ and V_T_ and percentage mean difference (%) for four PET radioligands. The mean, SD between subjects and SD within subjects for each radioligand were taken from test-retest studies on healthy subjects. NB: the variance is likely to be higher in a more heterogeneous clinical population, which will lead to smaller effect sizes for the same raw or % difference.

| Effect size | [<sup>11</sup>C]raclopride<br>BP<sub>ND</sub> (%) difference | [<sup>11</sup>C]AZ10419369<br>BP<sub>ND</sub> (%) difference | [<sup>11</sup>C]PBR28<br>V<sub>T</sub> (%) difference | [<sup>11</sup>C]UCB-J<br>V<sub>T</sub> (%) difference |
| --- | --- | --- | --- | --- |
| **Cohen’s D** |  |  |  |  |
| 0.2 | 0.07 (2%) | 0.03 (2%) | 0.37 (10%) | 0.36 (2%) |
| 0.5 | 0.17 (5%) | 0.07 (4%) | 0.92 (26%) | 0.9 (4%) |
| 0.8 | 0.27 (7%) | 0.12 (7%) | 1.48 (42%) | 1.43 (6%) |
| 1.2 | 0.41 (11%) | 0.18 (10%) | 2.22 (63%) | 2.15 (10%) |
| 1.5 | 0.51 (14%) | 0.22 (13%) | 2.77 (79%) | 2.69 (12%) |
| **Cohen’s Dz** |  |  |  |  |
| 0.2 | 0.03 (1%) | 0.03 (2%) | 0.12 (3%) | 0.21 (1%) |
| 0.5 | 0.08 (2%) | 0.07 (4%) | 0.3 (8%) | 0.53 (2%) |
| 0.8 | 0.13 (4%) | 0.11 (6%) | 0.48 (13%) | 0.85 (4%) |
| 1.2 | 0.19 (5%) | 0.16 (10%) | 0.71 (20%) | 1.28 (6%) |
| 1.5 | 0.24 (7%) | 0.2 (12%) | 0.89 (25%) | 1.6 (7%) |

### Objective 1 - keeping false positive rate below 5%

#### Simulation set-up

Our first aim of this tutorial is to compare sequential BF testing to the conventional way of evaluating research hypotheses in a clinical PET study, which is to apply a single test after the entire sample size has been collected (i.e., a “fixed N approach”).^6^ Here we will explore the differences between sequential BF testing and taking a fixed N approach, presenting the reader with the trade-offs that are made when using a sequential procedure. We will show how sequential BF testing can be used in order to stop a study early while still keeping the rate of false positive findings under the commonly set threshold of 5%. In order to compare the sequential BF framework to the conventional approach, we will only stop data collection when there is evidence in favour of an effect. In this simulation, evidence for the null hypothesis will hence be treated the same as inconclusive stopping outcomes, i.e. no early stopping decisions will be made when BF < 1. For a demonstration on how to stop a study early also when H_0_ is supported, see Objective 2 below.

Due to the high cost of performing a PET scan, it is rare to see PET studies with large sample sizes. In order to reflect this reality, our simulations focus on typical cases in PET, when the study includes between 12 to 100 subjects/group. Specifically, for the cross-sectional design we:

1. Simulated a population of patients and control subjects with a true difference between the groups corresponding to *δ*.
2. From each population we sampled one patient and one control and compared the difference using the default two-sample BF t-test from the R-package *BayesFactor* (Morey and Rouder 2018).
3. The data collection was stopped if the BF reached a predefined threshold.
4. If BF did not reach a predefined stopping threshold, we repeated steps 2-3 until we reached Nmax subjects/group. At that point, the data collection was stopped, regardless of the BF result.

Step 2-4 were then repeated 30,000 times, and the results from the sequential BF testing were saved. We examined a range of *δ* values, from a Cohen’s D = 0 (no difference between patients and controls) to a Cohen’s D = 1.5 (large difference between patients and controls). We also evaluated a range of stopping thresholds, going from 2 (negligible evidence for H_1_) to 10 (strong evidence for H_1_).

The same simulation scheme as above was used for the paired design, with the exception of simulating within-subject differences and applying a paired BF t-test instead of a two-sample test.

Sequential BF testing has been shown to produce errors, leading to wrong decisions, most often when sample sizes are very low (Schönbrodt et al. 2017). If the first BF is calculated soon after the initiation of the study (e.g., at n = 3 subjects/group), then the false positive rate can become unacceptably high. It is therefore sensible to first collect a fixed number of subjects from each group before initiating sequential testing with BF. Below we present the results from simulations within which sequential BF testing began after data from 12 subjects/group had been collected.

## Results

Figure 3A and 3B show the estimated false positive rate when applying sequential BF testing in the case in which there is no difference between the two groups. Three different stopping thresholds have been used (BF > 3, 4 and 6, respectively). When the maximum allowed sample size increases, the false positive rate goes up. This is because the longer the sequential testing can go on, the more decisions are being made. Some of these decisions will be wrong, meaning that the researcher will claim support for an effect, when in fact there is no effect in the population. Eventually, when N becomes high enough, the rate of false positives will reach an asymptote (see Supplementary Figure S1). But with a maximum sample size above 30 subjects/group, it is not possible to keep false positives below the 5% using a decision threshold of 4. Instead, if the researchers wish to control the false positives at the 5% level, one option is to increase the decision threshold. Figure 3C and 3D show the estimated decision threshold that would be needed to keep the false positive rate at 5% when using sequential testing with the default BF t-test, while still starting at 12 subjects/group and checking the BF after the collection of each patient-control pair.

**Figure 3:**
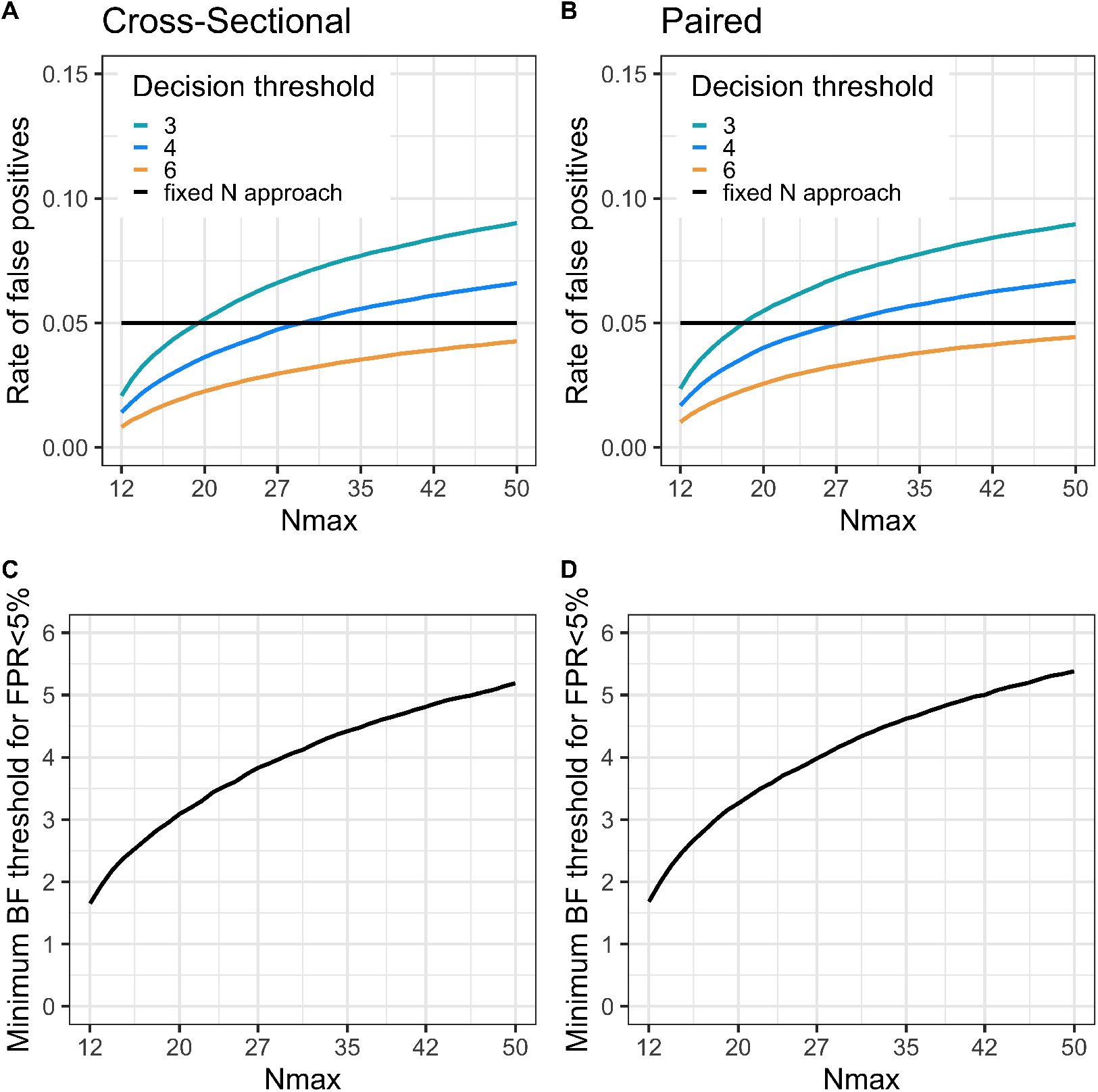
A and B) The rate of false positive stopping decisions increases as the maximal number of subjects per group (Nmax) becomes higher due to an increasing number of decisions being made, shown for three different BF decision thresholds. C and D) The BF decision threshold can be adjusted to achieve a desired rate of false positive evidence (here 5%) for different Nmax. For all figures: samples are drawn from populations showing no (between or within) mean difference; testing starts at N = 12 subjects/group; and BF is checked after every additional comparison pair (1 set of patient-control scans or pre-post scans). FPR = false positive rate.

Figure 4 shows the results from the simulations when Nmax=30 subjects/group, using a decision threshold just above 4. In the upper panel the rate of positive studies at different population effects is visualised. The curves cross the y-axis at D=0 (i.e., the false positive rate), 5% for both BF seq and the conventional fixed N test. As can be seen, had the researcher instead applied a fixed N approach (at 30 subjects/group), they would have increased the power to detect the population effect by no more than ∼10% for any effect size compared to using sequential testing. Note that when calibrating the BF threshold to correspond to a 5% false positive rate, and stopping only for H_1_, the BF procedure will show identical results to performing sequential NHST t-tests with a 5% corrected significance level.

**Figure 4:**
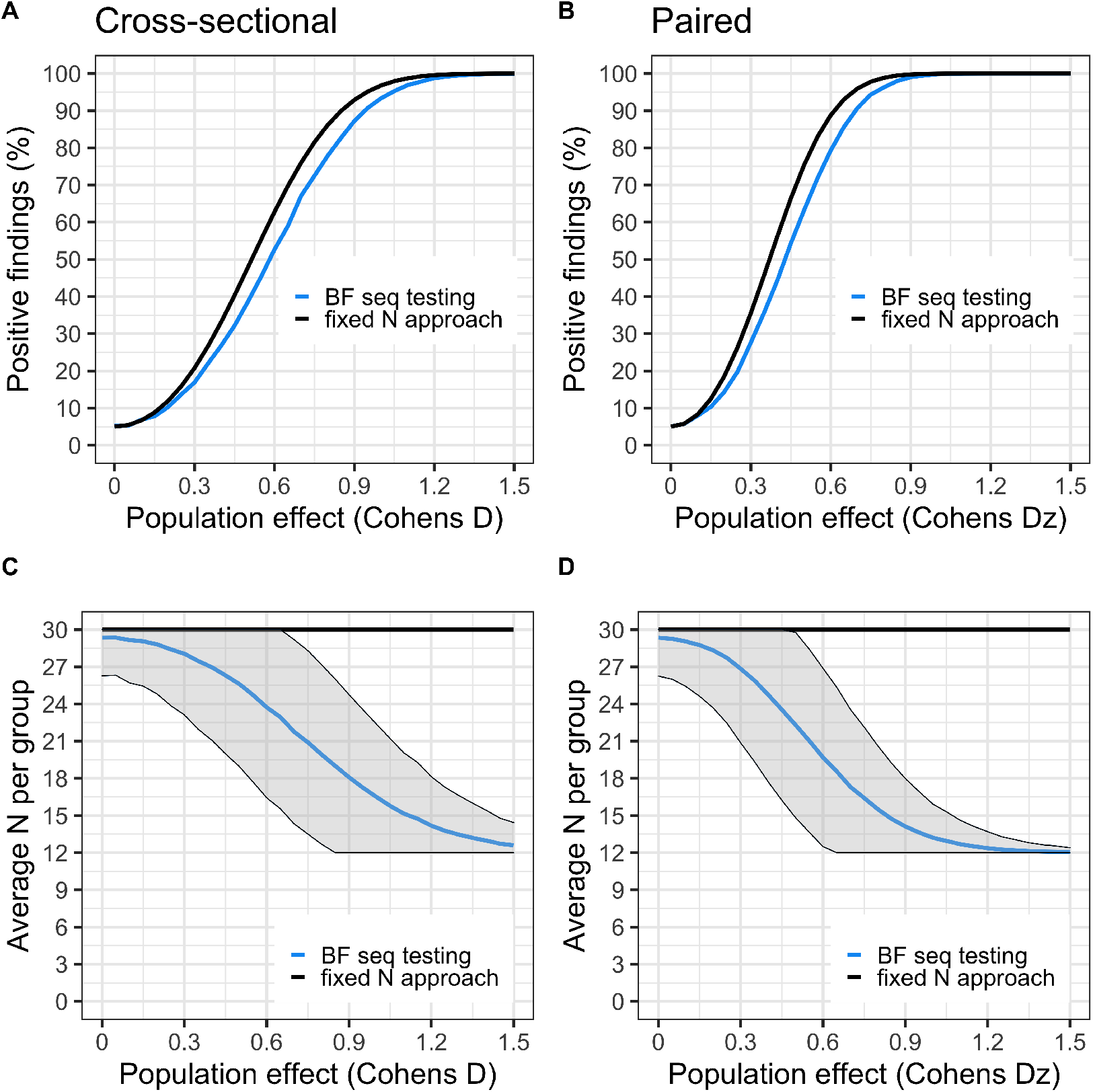
Panels A and B show the proportion of studies that showed support for H_1_ (aka “power curves”) for BF sequential testing (blue) and fixed N test (black), at different population effects (starting at D = 0, i.e. no effect). For the fixed N approach, only one test is performed at N = 30 subjects/group. For the sequential testing, 12 subjects/group are first collected, then BF is checked after each added comparison pair until 30 subjects/group is reached, stopping the study when BF > 4. Panels C and D show the average number of subjects needed to reach a stopping decision at different population effects. The fixed N test is the black line (N = 30 subjects/group); BF sequential testing is the blue line with shaded area denoting ± 1SD.

Figures 4C and 4D show the average number of subjects needed to reach a decision at different population effects. As the underlying difference between the two populations is increased, fewer subjects are needed to reach a decision using sequential BF testing. Already at a true population difference of Cohen’s Dz = 0.5, researchers will on average save 30% in terms of both expense and amount of injected radioactivity when applying sequential BF testing compared to the fixed N approach.

In summary, if the PET researchers apply a sequential testing procedure as presented here, they have the potential of reducing both expense and radioactivity exposure, but it comes at a cost of a lower rate of true positives (i.e., statistical power), for a range of effect sizes.

Simulation results for different choices of Nmax (15, 20, 50 and 100 subjects/group) are presented in supplementary information (Supplementary Figures S2-S5).

Practical recommendations 1

In this simulation set-up we show that it is possible to stop a PET study early if there is a true underlying effect in the population, while keeping the number of false positive findings below 5%. We recommend to not start sequential BF testing until at least 12 subjects per group have been collected, adjusting the BF threshold upwards for larger values of Nmax. Importantly, these settings should only be used when the PET researcher is interested in stopping early when data shows support for H_1_. All BF values showing support in favour of H_0_ (BF < 1) should not, with these settings, be interpreted as anything else but inconclusive stopping outcomes.

### Objective 2 - stopping for both H_1_ and H_0_

The second aim of this tutorial is to assess whether sequential BF testing can be used to stop a clinical PET study early, both when there is an effect in the population, but also when the population effect is zero. After the collection of each data point, we will compare both the BF, and its reciprocal, to an *a priori* set decision threshold. If either the BF or its reciprocal passes the threshold, the study will be stopped, and we will declare support in favour of the alternative or null hypothesis, respectively.

#### Simulation set-up

We used the same simulation set-up as above, where a grid of maximum sample sizes and effect sizes ranging from 0 to 1.5 was used to evaluate sequential BF testing in a cross-sectional and paired design respectively. We used the same settings for the default BF t-test as above with one exception: the alternative hypothesis was still a Cauchy distribution (centered at zero with r=0.707) but now specified as being one-sided, instead of two-sided. This means that we anticipate that the effect will go in one direction (e.g., patients will have a higher BP_ND_ than controls), making the test into a one-sided default BF t-test. The reason for considering only a one-sided test in this scenario is that it is not possible to stop for H_0_ when using a two-sided test at smaller sample sizes when using reasonable decision thresholds (BF < ¼, see Supplementary Figure S10).

In the simulations reported below, a one-sided default BF t-test was therefore performed sequentially, after 12 subjects per group had been collected. If no decision was reached after 30 subjects/group had been collected, the study was stopped and the result was considered to denote an inconclusive stopping outcome. The decision threshold was set to 4 and ¼, for the alternative and null hypothesis respectively.

## Results

Figure 5 summarises the results from the simulations. When the population difference is zero in the cross-sectional design, a study will stop (incorrectly) for H_1_ just above 5% of the time, and (correctly) for H_0_ about 60% of the time. In this case, studies will be able to stop, on average, after just 21 subjects/group have been scanned. Hence, assuming there is no true difference between groups, a sequential BF testing procedure will on average save 18 PET examinations, i.e., 30% in terms of expense and exposure, compared to the strategy of collecting data until Nmax is reached.

**Figure 5:**
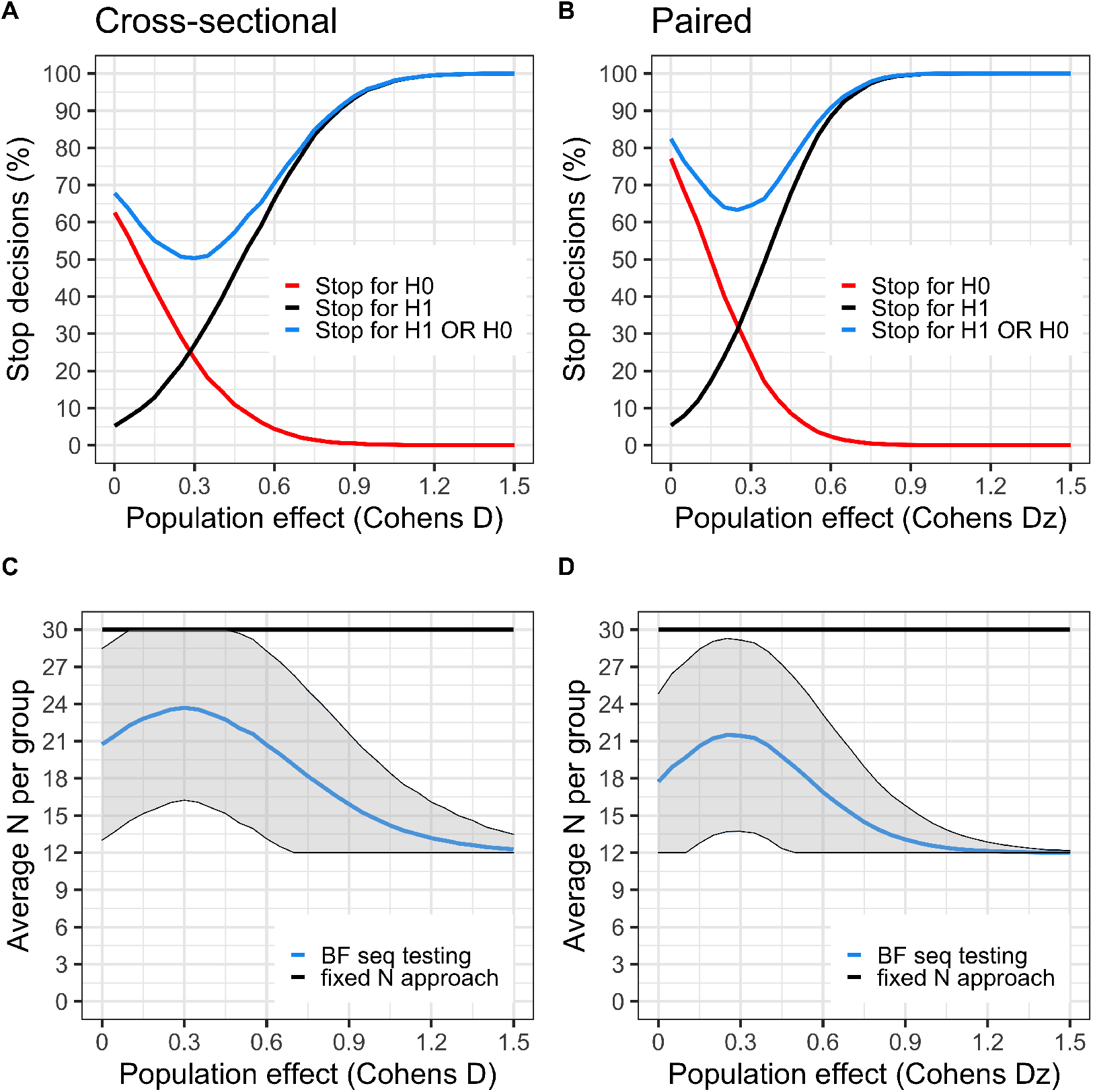
A and B) The black curve denotes the proportion of studies that showed support for H_1_ (BF > 4) during data collection, at a range of population effects (starting at no effect, D = 0). The red curve is the proportion of studies that showed support for H_0_ (BF < ¼). The blue curve is the sum of the red and black curves. It describes the proportion of studies that yield a stopping decision, either in favour of H_0_ or in favour of H_1_. Since the true population effect is unknown to the researcher, the blue curve can be viewed as the probability of reaching a stopping decision regardless of it being due to true or false evidence. When there is no effect in the population (D = 0), the black curve shows the rate of false positives. At this point, the red curve shows the probability of correctly stopping early for H_0_ (i.e., proportion of true negatives). C and D) shows the average number of subjects needed to reach a stopping decision at different population effects. The flat black line represents Nmax (30 subjects/group). BF sequential testing is the blue line with shaded area denoting ± 1SD.

For the paired design we observe a similar false positive rate but a higher rate of true negative evidence (∼75%) when the population effect is zero. The rate of true positives is also higher at all evaluated effect sizes compared to the cross-sectional design (Figure 5). For example, at a population effect of Dz=0.5, a stop decision is reached before or at Nmax in 75% of studies. In this case, studies can on average be stopped after scanning 19 subjects/group, saving in total 22 examinations, i.e., 36% expense and injected radioactivity.

The area around the point where the red and black line cross, is a weak spot for the BF sequential testing procedure. At this range of true effects in the population, the risk of obtaining an inconclusive stopping outcome is at its highest (i.e., the blue line is at its nadir). In the cases in which a decision is reached, the study will stop with equal probability for H_1_ and H_0_, and the risk for false negative evidence is around 25%.

Assuming that the PET researcher would be interested in stopping a study early when H_0_ is supported, a range of population effects with high risk of false negative evidence will always exist. It is therefore sensible to beforehand decide on a minimal population effect of interest, and choose the settings to ensure that any larger effect does not have too high a risk of generating evidence for H_0_. For example, with the settings presented in Figure 5 (Nstart = 12, Nmax = 30, threshold = 4) it could be said that all effects between D = 0 and 0.45 ought to be of little clinical interest, in order for sequential BF testing to be applicable in this case since the risk of false negative evidence for these effects is > 10% ^7^.

The same results but for different Nmax (15, 20, 50 and 100 subjects/group) are presented in supplementary information (Supplementary Figures S6-S9).

Practical recommendations 2

In this simulation set-up we show that it is possible to stop a PET study early either when there is a true underlying effect in the population, or when the effect is zero. When using the Cauchy distribution to describe the alternative hypothesis, we recommend that sequential BF testing does not start until at least 12 subjects per group have been collected, setting the BF threshold to a minimum of 3, and using a one-sided test. Using a one-sided test means that the researcher must make an *a priori* prediction of which direction the effect will have, and not change this prediction after the study is started, similar to performing a classical one-sided NHST t-test.

### Objective 3 - Application to real clinical PET data

The third and final aim of this tutorial is to apply sequential BF testing to a real clinical PET setting. To this purpose, we used already collected data of patients with major depressive disorder and healthy control subjects examined with [^11^C]WAY100635, a radioligand which binds to the serotonin 1A (5HT1A) receptor (Parsey et al. 2010; Chen et al. 2019). From this data, we included 40 medication-free patients (mean Age 36.2 (12.9SD); 25 females) and 40 healthy controls (mean Age 37.1 (14.0SD), 25 females), using BP_F_ (specific binding over the free-fraction in plasma) as the primary outcome measure for assessing 5HT1A receptor availability in the brain. The 5HT1A receptor acts inhibitory on serotonergic neurons in the raphe nuclei. A high concentration of receptors in raphe will likely lead to lower transmitter release in serotonergic projection areas. Hence, we hypothesized that patients suffering from MDD show higher 5HT1A receptor availability in the raphe nuclei compared to healthy controls. Here we apply sequential BF testing to examine whether this hypothesis gains enough support to stop data collection before 40 subjects per group are reached.

## Methods

We applied sequential BF testing to assess the support in data in favor of patients having higher BP_F_ in the raphe nuclei than controls (H_1_), as compared to no difference (H_0_). We also examined the evidence in data in favor of H_0_ over H_1_, to stop the study early if the null hypothesis was supported. The stopping threshold was set to BF>5 for H_1_ (BF < 1*/*5 for H_0_), in order to keep the rate of false positive evidence below 5%^8^ (see Supplementary Figure S1). We used a one-sided Cauchy distribution, centered around 0 with an *r* = 0.707 to describe the predicted mean difference under H_1_. H_0_ was specified as the point zero value.

First, patient and control data were sorted according to the chronological order in which the subjects were examined. We then retrieved the first 12 patients and 12 control subjects, standardized all raphe nuclei BP_F_ values, calculated the BF and compared it against the stopping thresholds. Following this, we added one additional patient and one healthy control to the previous subjects, checked BF against the thresholds, and so on, until we either were able to stop early, or reached the maximum of 40 subjects/group.

## Results

When applying a one-sided two-sample t-test to the full dataset, we observed a large group difference in raphe nuclei BP_F_ (t = 4.1, df = 76.58, p = 0.00006, BF = 385), with patients showing higher values compared to healthy controls (D = 0.91 or a 51% increase).

When applying sequential testing, the BF passes the threshold of 5 in support of H_1_, after the inclusion of 27 subjects/group (Figure 6). Hence, had sequential testing been applied in collection of [^11^C]WAY100635 patient-control data, it would have been possible to stop the recruitment at a total N of 54 instead of 80, saving 33% in terms of expense and radioactivity exposure. Assuming that a PET examination costs 5000 USD/Euro, that would amount to saving 130,000 USD/Euro in total.

**Figure 6:**
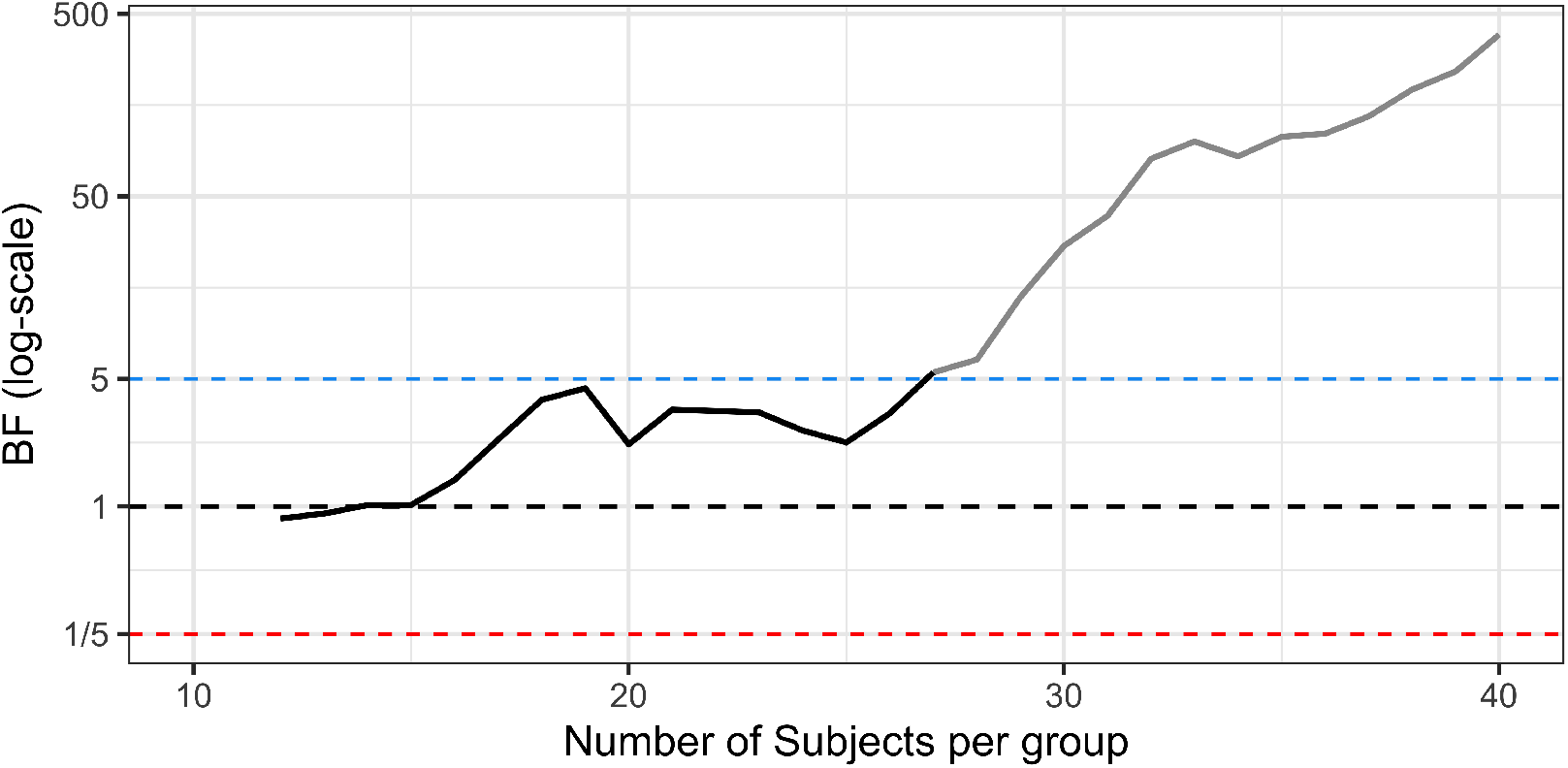
Sequential BF testing applied to real PET data. Patients with major depressive disorder were compared to healthy control subjects using [^11^ C]WAY100635 BP_F_ as a measure of 5HT1A receptor availability in the raphe nuclei. BF testing starts after 12 subjects/group have been collected and a stop decision is triggered at 27 subjects/group. The blue line denotes the stopping threshold for H_1_ (set to 5) and the red line the stopping threshold for H_0_ (set to 1/5). The black line shows the change in BF following the inclusion of each additional patient-control pair. The grey line shows how the BF trajectory would have looked, assuming no stopping decision was made, and the data collection instead continued up until the full sample of 40 subjects/group.

## Discussion

In this tutorial we show that it is possible to stop recruitment of subjects in PET studies as soon as enough data have been collected to make a conclusion, reducing both expense and exposure to radioactivity. We do this by employing sequential BF testing, which assesses the support in data in favor of two competing hypotheses while the study is still ongoing.

When applying sequential BF testing, and the true population effect is small, the relative savings are modest compared to using the conventional design where only one test is performed at a fixed sample size. As the population effect becomes bigger however, it is possible to (on average) stop the data collection early, potentially saving more than half of the resources that would be needed for a PET study carried out with the conventional approach. For a range of effects, a fixed N test will however show around 5-10% higher true positive rate compared to sequential BF testing (Figure 4A-B). By using a lower BF decision threshold, the proportion of true positive stopping decisions will increase when there is an effect, but it comes with a cost of more false positives when H_0_ is true.

### Alternative sequential procedures

There are sequential testing methods other than BF that can be used to stop the data collection early, and that offer exact control over error rates. For the NHST framework, a viable alternative is to apply a so-called group-sequential NHST design (Schulz and Grimes 2005). In fact, when calibrating the BF threshold (using a “default” Cauchy-distribution for describing H_1_) to a 5% false positive rate when stopping only for H_1_ (see Objective 1), the sequential BF procedure will be identical to performing corrected sequential NHST t-tests using so called Pocock boundries. For NHST, there are however several different efficient strategies for doing sequential testing to stop a study as soon as H_0_ can be rejected (Proschan et al. 2006; Lakens 2014). One alternative is to use “alpha-spending functions,” where the significance threshold changes for each sequential test, making it harder to reject H_0_ at an early stage of the study, but easier later on, while still keeping the total false positive rate below 5% (Demets and Lan 1994; Selwyn and Fish 2004). There are also methods for performing so called “futility testing,” with which the researchers can stop a study early if it is deemed to unlikely that a significant finding will be achieved later on (Lachin 2005). Addressing such sequential-NHST procedures for application in clinical PET research are however outside the scope of this tutorial.

### Evidence in favor of H_0_

Sequential BF testing allows a PET study to be stopped also when there is evidence in favor of no effect in the population, i.e., when H_0_ is true. If the researcher wants to use a sequential BF to assess support for both H_1_ and H_0_, we recommend using a one-tailed default BF t-test. This entails that the researcher must make an *a priori* prediction on the direction of the effect (e.g., patients have higher binding than control subjects, or vice versa). Without using one-sided default BF t-test, it will be difficult (or even impossible) to stop a study early in support for H_0_, given the sample sizes used in typical PET studies (N<100 subjects/group) while still using a threshold that is high enough to keep the rate of false evidence at reasonable levels (see Supplementary Figure S11).

When allowing a study to be stopped early for H_0_, false negative evidence must be considered as an additional type of error, and PET researchers now have to consider keeping both false positive and negative evidence low. To do so, the researcher has three main options to their disposal:

1. Increase the *a priori* set decision threshold for stopping (see Figure 3).
2. Recruit and scan a larger pool of subjects (>12/group) before initiating the sequential BF testing.
3. Check BF less frequently, e.g., only after the collection of each second or third (etc.) comparison pair, instead of after each single one. These three approaches could be used separately or in combination with each other. They do all however trade off against a higher number of included subjects, meaning that the study will, on average, require more subjects before a stop decision can be made.

When stopping also for H_0_, it is still possible to keep the rate of false positive evidence at or below 5% by increasing the decision threshold with higher values of Nmax (see Supplementary Figure S1). However, in order to keep the false negative evidence rate within reasonable limits, we do not recommend setting the decision threshold lower than 3, regardless of Nmax.

Figure 5 presents the true and false evidence rates the researchers can expect when using a one-sided BF t-test and when they are correct in their prediction of the direction of an effect. If there is a sizable patient-control difference in the population, but it goes in the opposite direction to the predicted one, then using this approach would lead studies to be stopped early due to the null-hypothesis being supported almost 100% of the time. Of course, the null is not true in this case, but it is closer to the true population effect than the *a priori* defined alternative hypothesis. Hence, it will often gain the most support from the data.

When allowing stopping for H_0_ as well, the researcher should be aware that the false negative evidence rate can become large when the effect is close to zero, but not exactly zero. Using sequential BF testing, researchers will run the risk of stopping the study, claiming support for no effect, when there is in fact a true but small effect in the population. Whether or not this is acceptable to the researchers depends on how small an effect has to be, in order for the researcher to consider it practically indistinguishable from zero. Using the sequential BF approach with the settings described in Figure 5 (e.g., a decision threshold of 4), the false negative evidence rate becomes high (>10%) for all effects between 0 and 0.45 for a cross-sectional design. Hence, if PET researchers consider this range of effects to be practically or clinically relevant, caution is warranted before applying a sequential BF testing procedure as shown here. Instead the researchers could increase the decision threshold, or specify H_1_ to assign larger plausibility to smaller effects, by lowering the width of the Cauchy (setting *r* to, e.g., 0.5) when planning the study. The latter will increase the rate of true positive evidence at smaller effect sizes, but it will also increase the rate of false positive evidence when H_0_ is true.

Example 3

There is a true difference in [^11^C]raclopride BP_ND_ between a patient and a control population that is of size Cohen’s D = 0.2 (a difference of 0.07 BP_ND_ or 2%, Table 2). For such a small effect, studies will often be stopped because H_0_ is supported when applying the sequential BF testing procedure, e.g., when using a threshold of 4 and ¼, respectively. A PET researcher might consider such an effect to be practically indistinguishable from zero. In such a scenario, support for H_0_ is hence of little concern, or even a preferred outcome for the researcher. On the other hand, if the researcher considers a Cohen’s D = 0.2 to be clinically relevant, then a sequential BF procedure using the default BF t-test cannot reliably give the correct stopping decision for H_1_ in commonly seen sample sizes in PET research (<100 subjects/group). If the researcher wants to be able to detect such a small effect, they need to use a different specification of H_1_, or a higher decision threshold (e.g. BF>10) together with a much larger Nmax.

### Inconclusive stopping outcomes

For any given PET-study where sequential BF testing is applied there will be a chance of reaching Nmax without crossing the pre-set decision threshold. In such cases, where the researcher ends up with an “inconclusive” stop decision, the BF is still interpretable. The suggested evidence thresholds in Table 1 can be used to report the support in data in favor of the two competing hypotheses, even though BF never formally reached the *a priori* set stopping threshold. For example, if the BF is above 3 (or below 1*/*3), this can still be reported as “moderate” support in data for one hypothesis over the other. However, in doing so the researcher cannot any longer say that they are controlling the rate of false evidence at a prespecified level.

### General considerations

By using a directional hypothesis (one-sided test) when applying BF sequential testing it is possible to stop a study at low sample sizes, when the null-hypothesis is supported. If the researcher wishes to allow stopping for H_0_, but uses a two-tailed default BF t-test instead, the average N will be much higher when using a reasonable BF decision threshold (>3, see Supplementary Figure S11).

Throughout this article we have used a Cauchy distribution with an *r* of 0.707 to describe the alternative hypothesis. However, for any given study design a more appropriate choice of H_1_ might exist. If a distribution with a higher *r* (e.g., 1) is used to define H_1_, then the researcher assigns more predictive weight to a mean-difference further away from 0. Doing so makes it easier to obtain evidence in favor of H_0_ (when there is no effect), but harder to obtain evidence for H_1_ when the true effect is small. This means that in using a larger *r* than 0.707 (all other settings the same); 1) the rate of false positive evidence will be lower; 2) the researcher will on average stop earlier when H_0_ is true; 3) small but true effects will more often produce false negative evidence (but see example box 3).

Increasing the width of the Cauchy can be desirable in e.g., a paired study design, where the variance can be expected to be low, but the difference in raw scores is assumed to be similar to that from a cross-sectional design. The reason for this is that the same change in raw score will correspond to a much larger standardized effect size (see Table 2), and it is therefore sensible to specify an H_1_ which predicts a more extreme difference. The lower rate of false positives that results from using a higher *r* (e.g., 1 instead of 0.707) can be utilized by starting the sequential BF testing earlier than at 12 subjects/group. As a result, the average sample size needed to reach a decision can be further decreased (see Supplementary Figure S10).

Practical recommendations 3

If the researcher has an informed idea on how the studied effect will look, they should consider using specification of H_1_ other than the “default” zero-centered Cauchy employed in this tutorial. This could e.g., be a normal distribution centered around a value considered to be either a plausible group difference or is clinically meaningful. This recommendation also applies to the null-hypothesis; we encourage the researcher to consider redefining H_0_ in order to test (what might be) a more appropriate hypothesis. An example of this could be to assess if an increase in patient BP_ND_ gains more support in data compared to a decrease, rather than just testing “there is a difference” vs. “there is no difference”.

In addition to the settings presented in this tutorial, there are several modifications that can be made for the BF test. For example, different decision thresholds can be applied for H_1_ and H_0_ depending on whether the researcher thinks that false positives are more important to avoid than false negatives, or vice versa. The researchers might also think, based on previous literature or bounds of a minimal effect size of interest, that a different specification of the alternative-hypothesis is more suitable to their research question (see “Practical Recommendations 3”).

This tutorial makes the assumption that, for a cross-sectional design, a scanned patient is always followed by a scanned control subject, or vice versa. In a real PET study, this is not always a feasible recruitment scheme. The results above also assume that PET researchers base their stopping decision on the outcome from one primary region of interest. If a researcher applied sequential BF testing, but cherry-picks the outcome from two or more regions to make a stopping decision, the risk for obtaining false evidence will increase. Finally, this tutorial only applies to study designs where a two-sample or paired t-test are suitable, and the reader cannot assume that the results would be similar if e.g. a regression model with covariates were to be used instead.

If PET researchers wish to use settings not discussed in depth in this tutorial: e.g., custom specification of H_1_, different decision thresholds for H_1_ and H_0_, other recruitment schemes, and/or statistical models, we recommend that they modify our simulation code to evaluate their own study design before starting the data collection. All code can be found freely available on https://github.com/pontusps/Early_stopping_in_PET.

### Specific recommendations

Before applying the sequential BF testing method, the researchers should decide what they want to prioritize: keeping the risk of making the wrong stopping decision low but accepting more inconclusive stopping outcomes, or stopping as early as possible but with a higher risk of errors. To help decide on this trade-off when planning PET studies, we have developed R functions with which PET researchers can examine how different settings for the sequential testing approach affects the average sample size needed, as well as rates of true and false evidence (https://github.com/pontusps/Early_stopping_in_PET).

It should by now be clear to the reader that a set of critical decisions needs to be made before applying sequential BF testing in a clinical PET study. For this reason, we recommend all researchers to pre-register their analysis before data collection starts^9^ (van ‘t Veer and Giner-Sorolla 2016; Poldrack et al. 2017; Knudsen et al. 2020). A pre-registration can be thought of as a safety net for the researcher. It helps guide the analysis and interpretation of data so that error rates are kept under control. It can also be shown to reviewers or readers to increase the credibility of the methods and findings. Deviations from a pre-registration is of course possible, and often warranted, but should be reported transparently in the article.

If a PET researcher wants to perform a study using sequential BF testing, we recommend that they follow the steps outlined in a flow-chart found in supplementary information (Supplementary Figure S12). In order to perform the default BF t-test, the freely available *BayesFactor* package (Morey and Rouder 2018) in R or point-and-click software JASP (Quintana and Williams 2018; Doorn et al. 2019; Love et al. 2019) can be used.

### Caveats

While a sequential testing procedure often allows the researcher to stop a study early, an important caveat is that the estimated effect size can become upwards biased. If a study can be stopped early due to H_1_ being supported, it is more likely that the observed effect size is above, rather than below, the true population effect (Schulz and Grimes 2005; Schönbrodt et al. 2017). This caveat should be considered before interpreting the effect size from a PET study that was stopped early, or before entering such an effect size into a meta-analysis.

### Further reading

If the reader is interested in learning more about sequential testing the following articles are a good start for a BF approach: Rouder (2014), Schönbrodt et al. (2017) and Schönbrodt and Wagenmakers (2018); and for a NHST approach: Schulz and Grimes (2005), Proschan et al. (2006) and Lakens (2014).

## Data Availability

The data code for reproducing the results, tables and figures in this article can be found at https://github.com/pontusps/Early_stopping_in_PET.

## Acknowledgements

We would like to thank Granville J. Matheson for valuable feedback on this project. PPS was supported by the Swedish Society for Medical Research and the Lundbeck Foundation.

## Footnotes

1 This range assumes that the injected dose of commonly used [^11^C]-isotope radioligands are between 100-850 MBq, with an average dose factor of 5.9 *µ*Sv/MBq.

2 The NHST approach is an amalgamation of Fisher’s and Neyman-Pearson’s theories of testing research data, and is not without criticism and controversy. However, a full outline of the procedure is beyond the scope of this paper and we refer interested readers to an excellent review by Perezgonzalez (Perezgonzalez 2015).

3 The Bayes Factor is also known as the “predictive updating factor” of Bayes Theorem. It is what updates the prior odds of two competing hypotheses into posterior odds after seeing the observed data, and hence describes the relative “predictive adequacy” of the hypotheses. Assuming that H_0_ and H_1_ have equal prior probability (P(H_1_) = P(H_0_) = 0.5), then a BF of 3 means that after conducting the study, H_1_ is 3 times more likely than H_0_. This corresponds to a posterior probability of 75% for H_1_ and 25% for H_0_ (Kass and Raftery 1995). Although posterior odds are the natural extension of the BF, researchers tend to only report the BF as a stand-alone metric for assessing different hypotheses, due to the inherent subjective nature of specifying prior odds.

4 It is worth emphasizing that, under reasonable assumptions, a Bayes Factor of 3 (three times more evidence in favor of the alternative-hypothesis over the null) corresponds to a p-value of approximately 0.05 – the most commonly set threshold for significance. A common misconception is that a p-value of 0.05 implies stronger support in data for the alternative. But at most times, it doesn’t (see e.g., https://www.shinyapps.org/apps/vs-mpr/ for more details). Due to this, a more strict threshold for declaring findings to be “significant” has been suggested in the statistical literature (Benjamin et al. 2017).

5 For the reader who is new to the BF, it might look peculiar to select an alternative hypothesis that not only includes H_0_ (point zero) but also places the highest probability mass at that point (see Figure 1). The alternative hypothesis in this case ought however to be interpreted as expressing the predicted plausibility of observing an effect in a range of values, rather than single points. This Cauchy distribution says that the researcher, before seeing the data, finds values in the range just around zero to be more likely than values far away from zero. There is nothing stopping the researchers from moving the center of the distribution away from zero when specifying H_1_, but that is beyond the scope of this tutorial.

6 Although a fixed N NHST is by far the most commonly used statistical paradigm in PET-research, the principles we demonstrate here will be true when comparing sequential testing to a fixed N BF t-test as well.

7 We believe that it is reasonable to consider a false negative to be somewhat less severe than a false positive, and for this reason we choose to present a 10% threshold here. However, just as with the conventional 5% threshold for false positive findings (Lakens et al. 2018), this is not a sacrosanct value. We encourage the reader to think for themselves what proportion of false evidence they are ready to accept when planning their own clinical PET studies.

8 A H_0_ stopping threshold of BF <^1/5^ means that if there is a true population difference between patients and controls in the range of D = 0 to 0.35, the risk of a false negative finding will be >10%. Hence, if the researchers consider such effects to be of high clinical interest to detect, they might consider using other settings instead. See the discussion for possible solutions.

9 See e.g., https://aspredicted.org/ for easy and efficient pre-registration.

