## Supplemental information for "Early stopping in clinical PET studies: how to reduce expense and exposure"

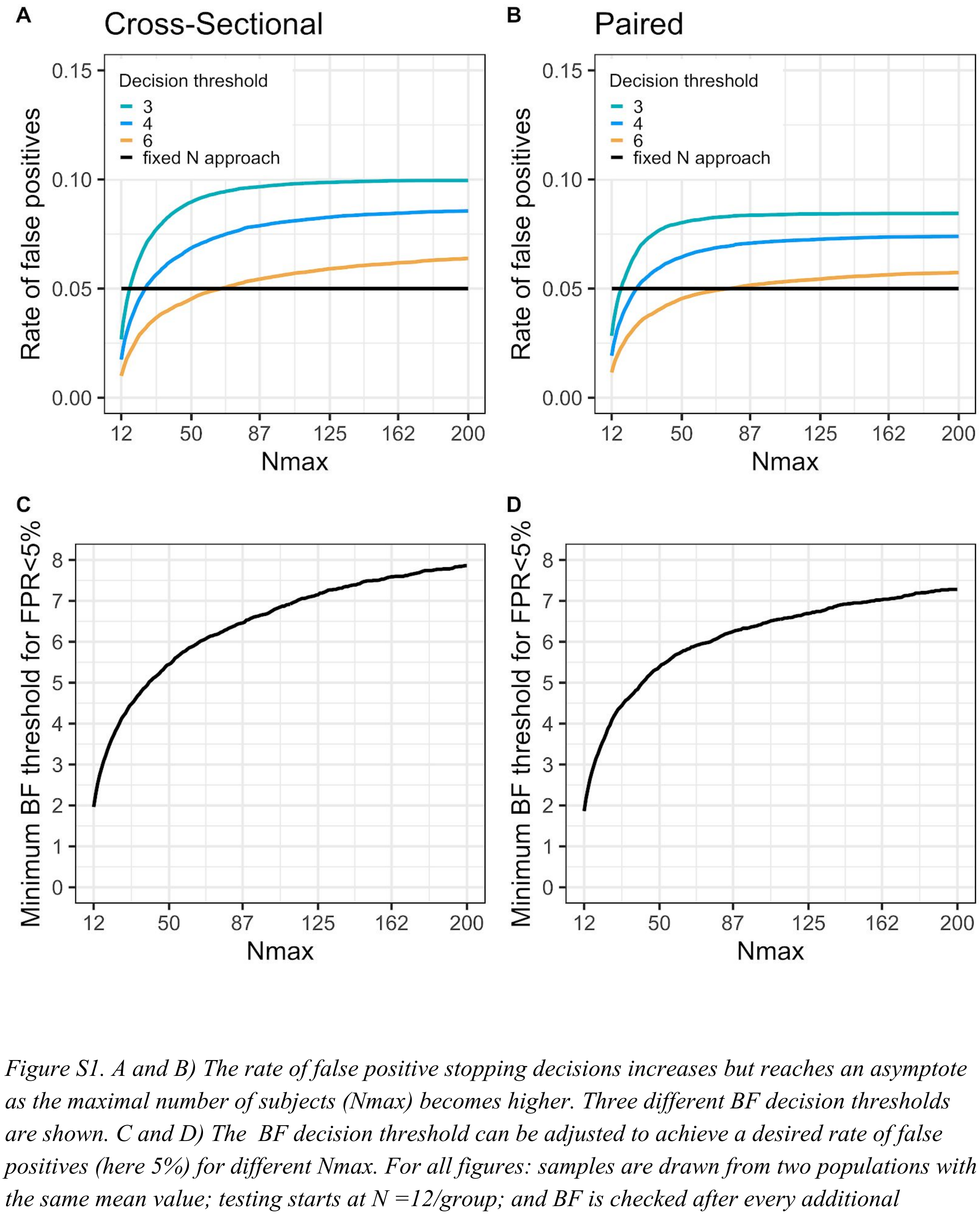


*Figure S1. A and B) The rate of false positive stopping decisions increases but reaches an asymptote as the maximal number of subjects (Nmax) becomes higher. Three different BF decision thresholds are shown. C and D) The BF decision threshold can be adjusted to achieve a desired rate of false positives (here 5%) for different Nmax. For all figures: samples are drawn from two populations with the same mean value; testing starts at N =12/group; and BF is checked after every additional comparison pair (1 set of patient-control scans or pre-post scans). Here stop decisions for H0 are allowed and the tests are one-sided.*

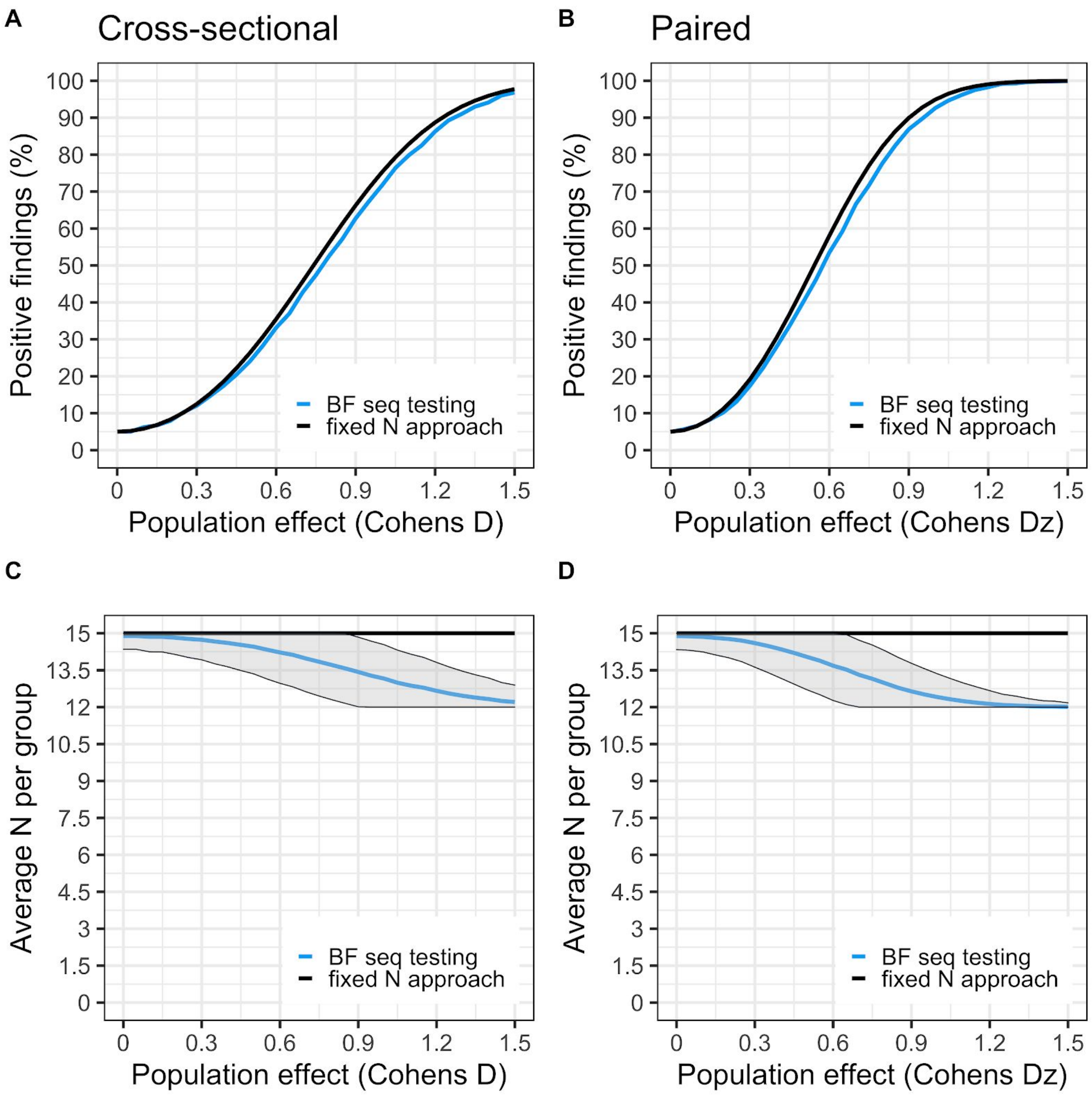


*Figure S2. Settings for the simulation: H1 is a two-sided cauchy(0,0.707), (Nstart = 12, Nmax = 15, BF threshold = 2.4 and 2.5 for cross-sectional and paired respectively). Panel A and B shows true positive (or "power") curves for BF sequential testing (blue) and a fixed N approach (black). The curves denote the rate of true positive findings at different population effects. For the fixed N approach, only one test is performed at N=15 per group. For the sequential testing, 8 subjects/group are first collected, then BF is checked after each added comparison pair until 15 subjects/group is reached, using a stopping threshold of 4. Panel C and D shows the average number of subjects needed to reach a stopping decision at different population effects. Fixed N is the black line (fixed at N = 15/group); BF sequential testing is the blue line with shaded area denoting ± 1 SD.*

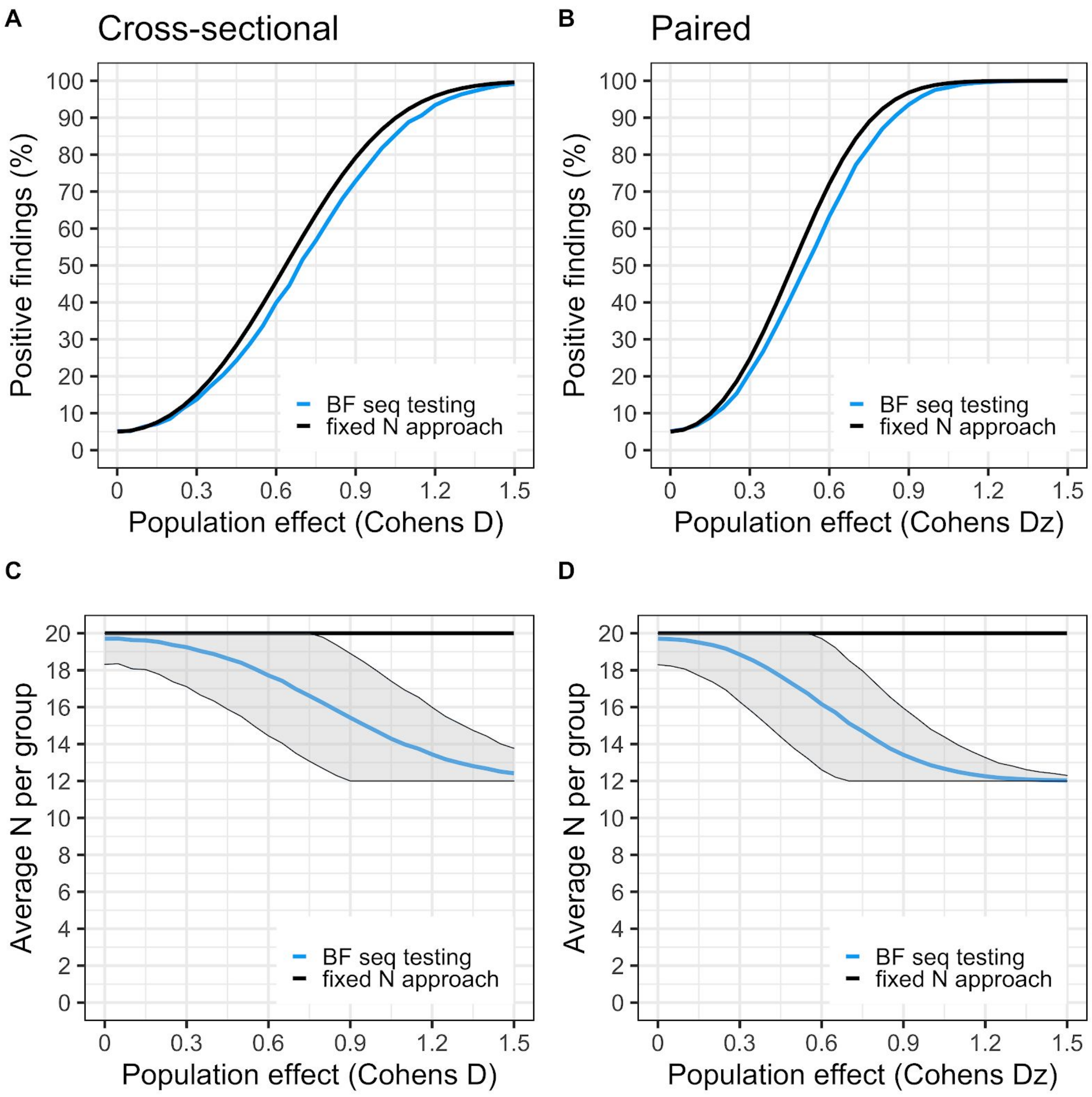


*Figure S3. Settings for the simulation: H1 is a two-sided cauchy(0,0.707), (Nstart = 12, Nmax = 20, BF threshold = 3.1 and 3.3 for cross-sectional and paired respectively). Panel A and B shows true positive (or "power") curves for BF sequential testing (blue) and a fixed N approach (black). The curves denote the rate of true positive findings at different population effects. For the fixed N approach, only one test is performed at N=20 per group. For the sequential testing, 12 subjects/group are first collected, then BF is checked after each added comparison pair until 20 subjects/group is reached, using a stopping threshold of 4. Panel C and D shows the average number of subjects needed to reach a stopping decision at different population effects. Fixed N is the black line (fixed at N = 20/group); BF sequential testing is the blue line with shaded area denoting ± 1 SD.*

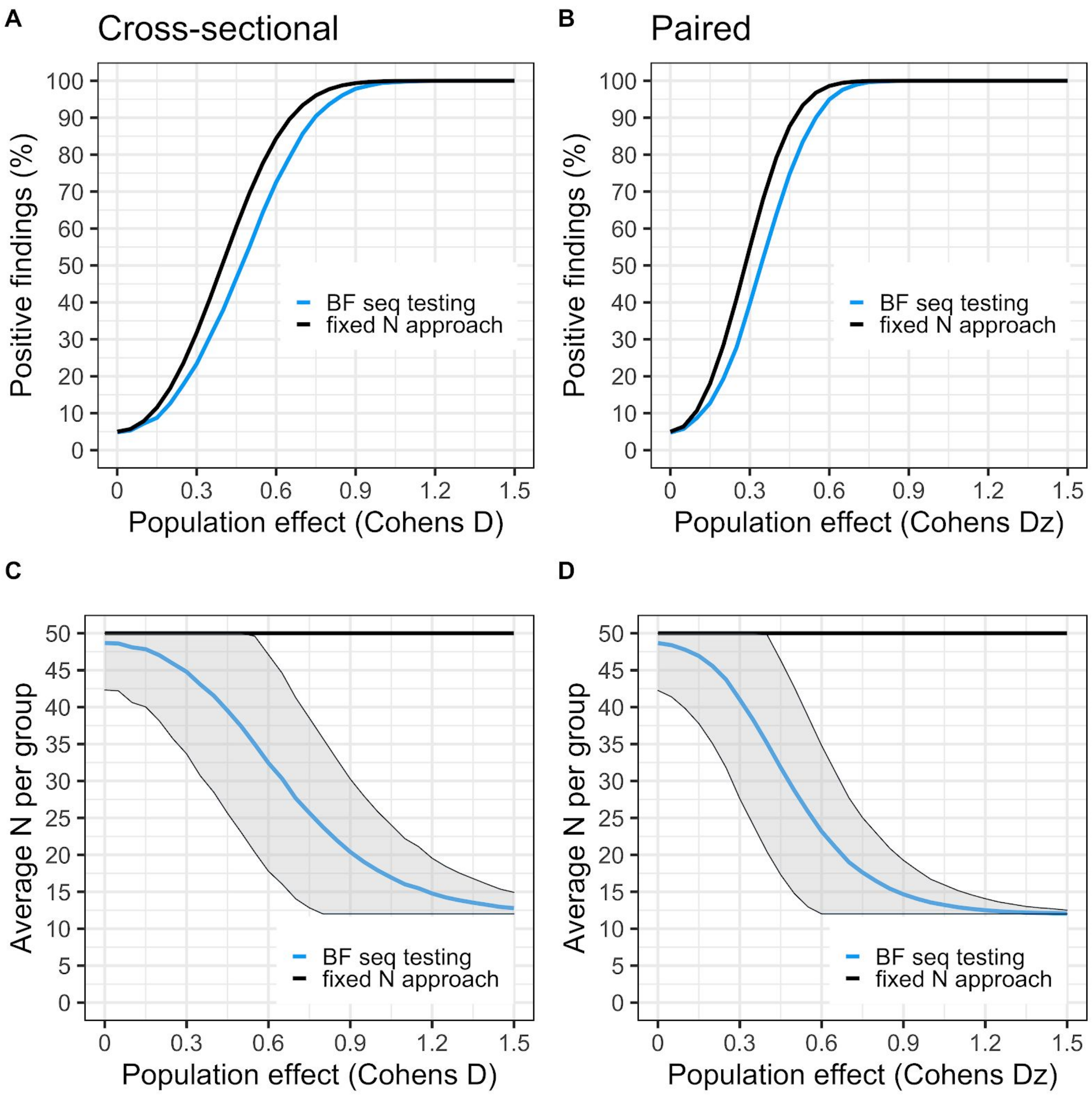


*Figure S4. Settings for the simulation: H1 is a two-sided cauchy(0,0.707), (Nstart = 12, Nmax = 50, BF threshold = 5.2 and 5.4 for cross-sectional and paired respectively). Panel A and B shows true positive (or "power") curves for BF sequential testing (blue) and a fixed N approach (black). The curves denote the rate of true positive findings at different population effects. For the fixed N approach, only one test is performed at N=50 per group. For the sequential testing, 12 subjects/group are first collected, then BF is checked after each added comparison pair until 50 subjects/group is reached, using a stopping threshold of 4. Panel C and D shows the average number of subjects needed to reach a stopping decision at different population effects. Fixed N is the black line (fixed at N = 50/group); BF sequential testing is the blue line with shaded area denoting ± 1 SD.*

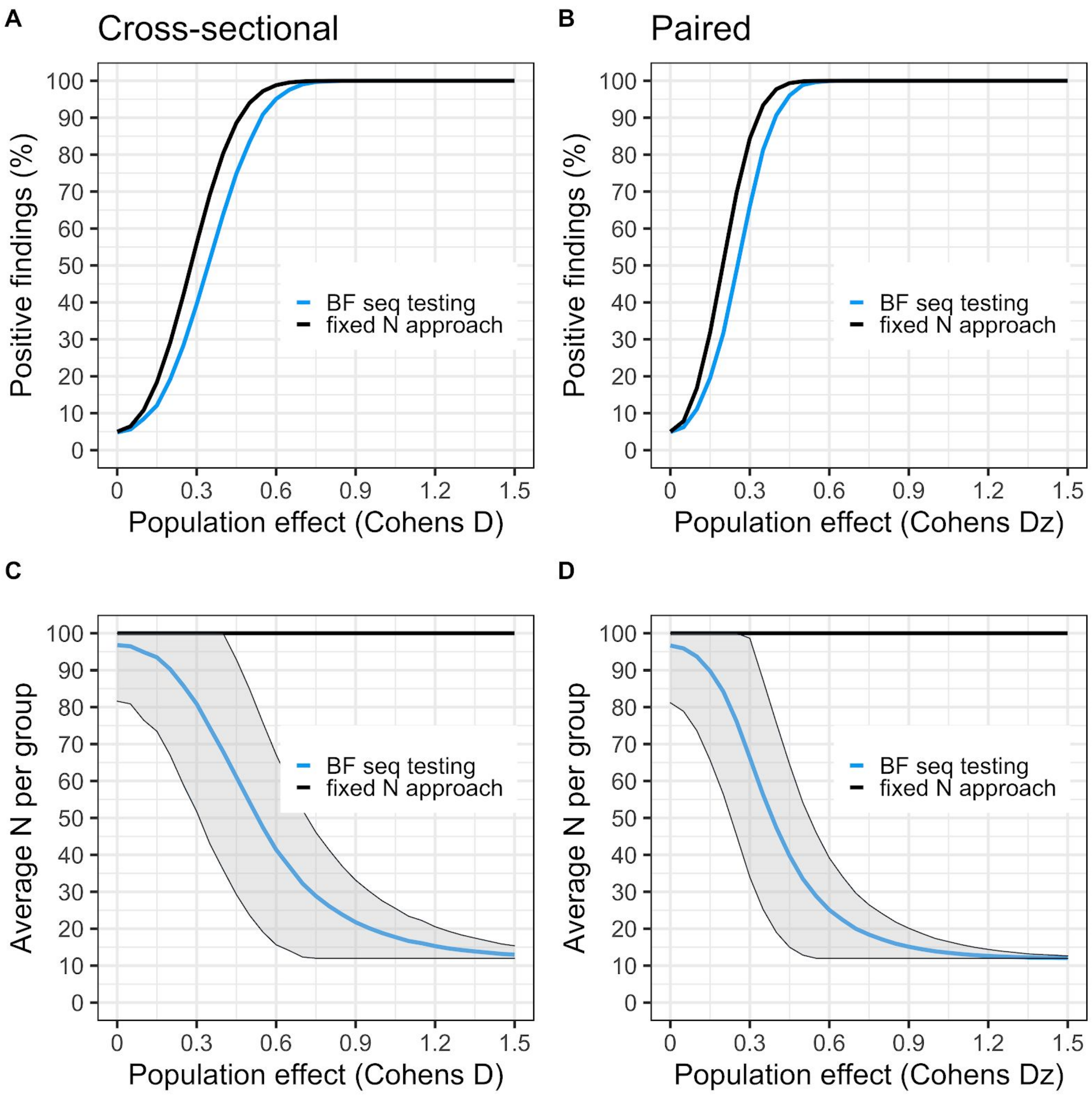


*Figure S5. Settings for the simulation: H1 is a two-sided cauchy(0,0.707), (Nstart = 12, Nmax = 100, BF threshold = 6.6 and 6.7 for cross-sectional and paired respectively). Panel A and B shows true positive (or "power") curves for BF sequential testing (blue) and fixed N approach (black). The curves denote the rate of true positive findings at different population effects. For the fixed N approach, only one test is performed at N=100 per group. For the sequential testing, 12 subjects/group are first collected, then BF is checked after each added comparison pair until 100 subjects/group is reached, using a stopping threshold of 4. Panel C and D shows the average number of subjects needed to reach a stopping decision at different population effects. Fixed N is the black line (fixed at N = 100/group); BF sequential testing is the blue line with shaded area denoting ± 1 SD.*

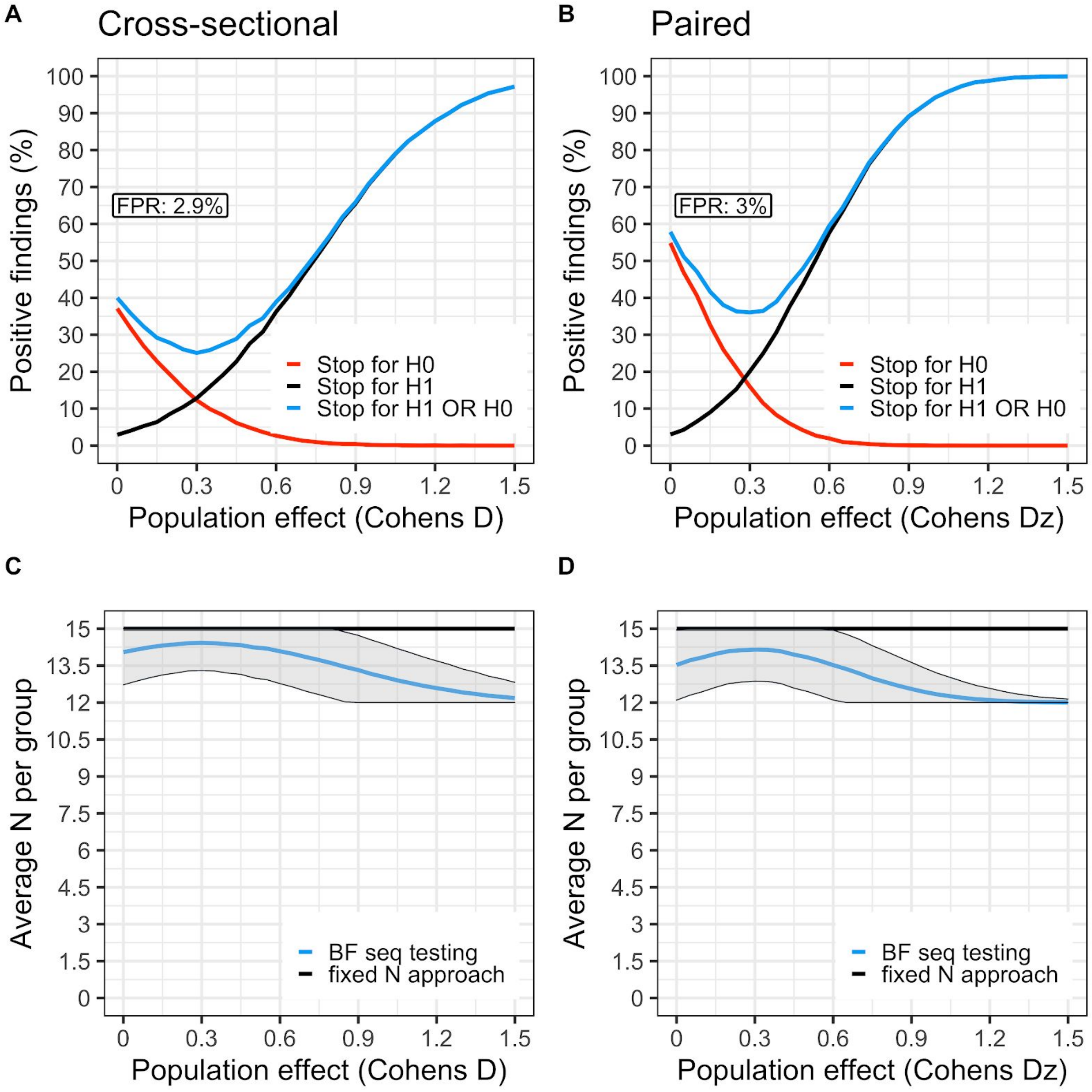


*Figure S6. Settings for the simulation: H1 is a one sided cauchy(0,0.707), (Nstart = 12, Nmax = 15, BF threshold = 4). A and B) The black curve shows the proportion of studies that showed support for H1 (BF>4) during data collection, at a range of population effects (starting at no effect, D = 0). The red curve is the proportion of studies showing support for H0 (BF<¼). The blue curve is the sum of the red and black curves. C and D) shows the average number of subjects needed to reach a stopping decision at different population effects. The flat black line represents Nmax (15 subjects/group). BF sequential testing is the blue line with shaded area denoting ± 1 SD.*

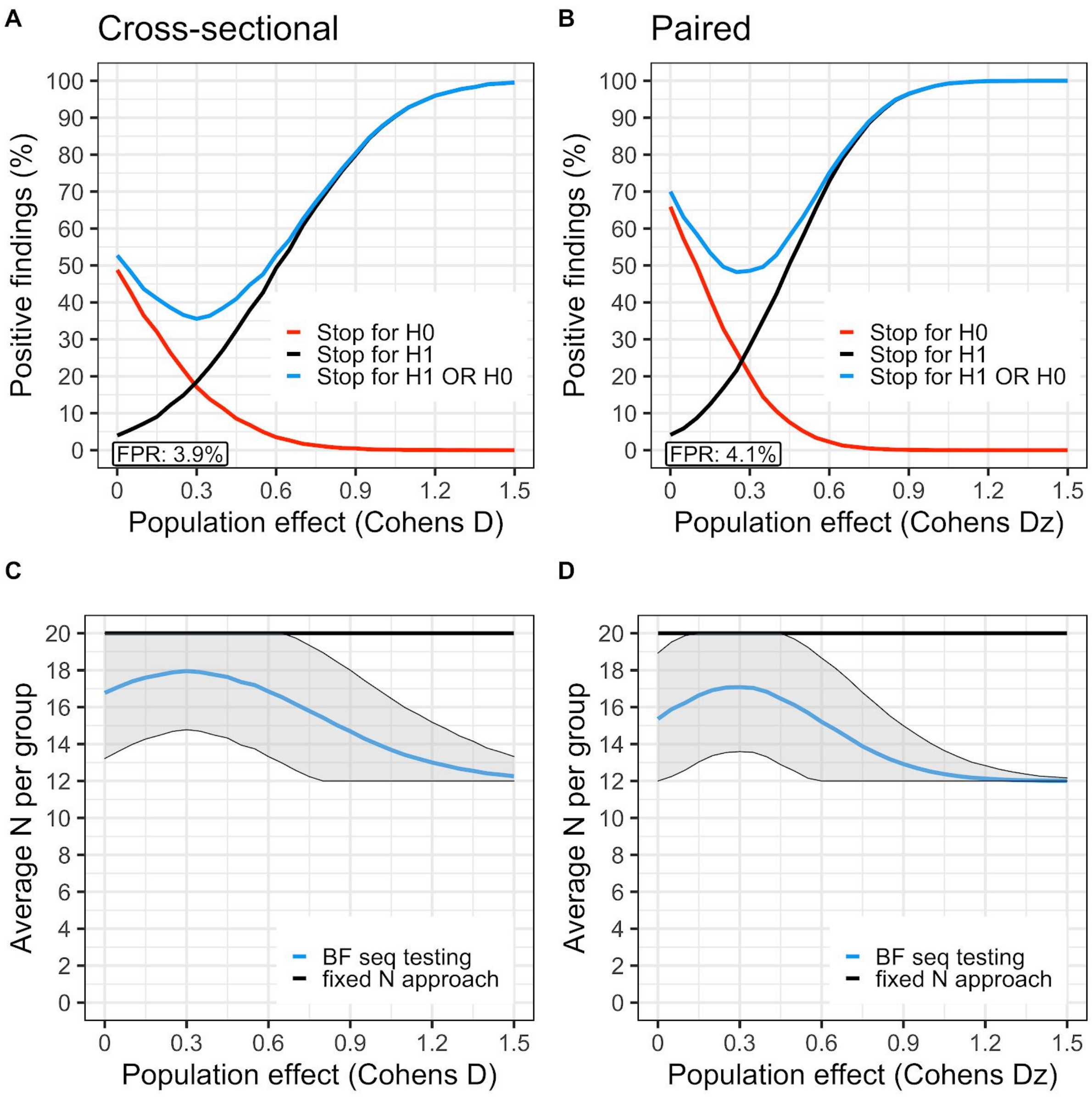


*Figure S7. Settings for the simulation: H1 is a one sided cauchy(0,0.707), (Nstart = 12, Nmax = 20, BF threshold = 4). A and B) The black curve shows the proportion of studies that showed support for H1 (BF>4) during data collection, at a range of population effects (starting at no effect, D = 0). The red curve is the proportion of studies showing support for H0 (BF<¼). The blue curve is the sum of the red and black curves. C and D) shows the average number of subjects needed to reach a stopping decision at different population effects. The flat black line represents Nmax (20 subjects/group). BF sequential testing is the blue line with shaded area denoting ± 1 SD.*

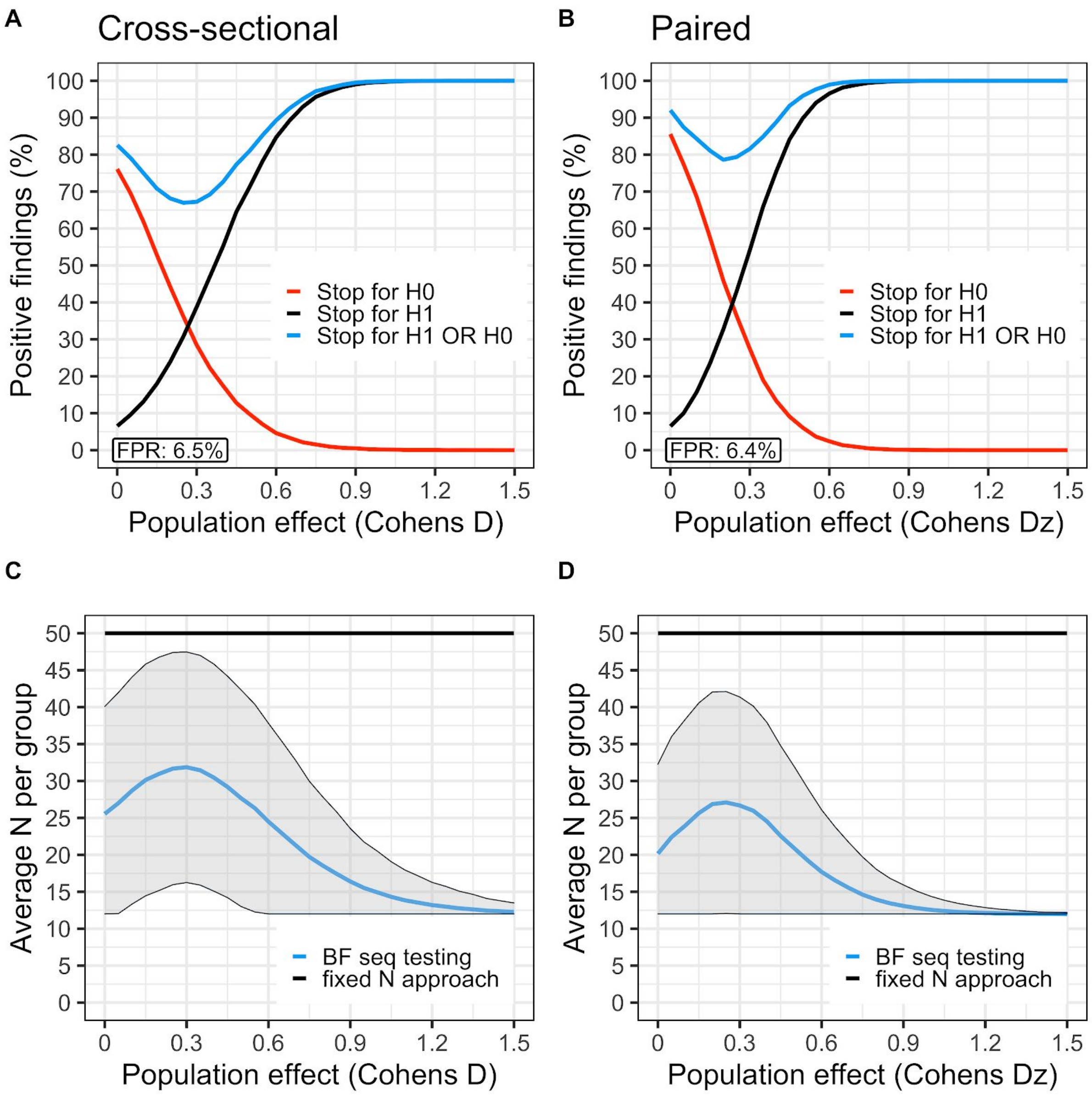


*Figure S8. Settings for the simulation: H1 is a one sided cauchy(0,0.707), (Nstart = 12, Nmax = 50, BF threshold = 4). A and B) The black curve shows the proportion of studies that showed support for H1 (BF>4) during data collection, at a range of population effects (starting at no effect, D = 0). The red curve is the proportion of studies showing support for H0 (BF<¼). The blue curve is the sum of the red and black curves. C and D) shows the average number of subjects needed to reach a stopping decision at different population effects. The flat black line represents Nmax (50 subjects/group). BF sequential testing is the blue line with shaded area denoting ± 1 SD.*

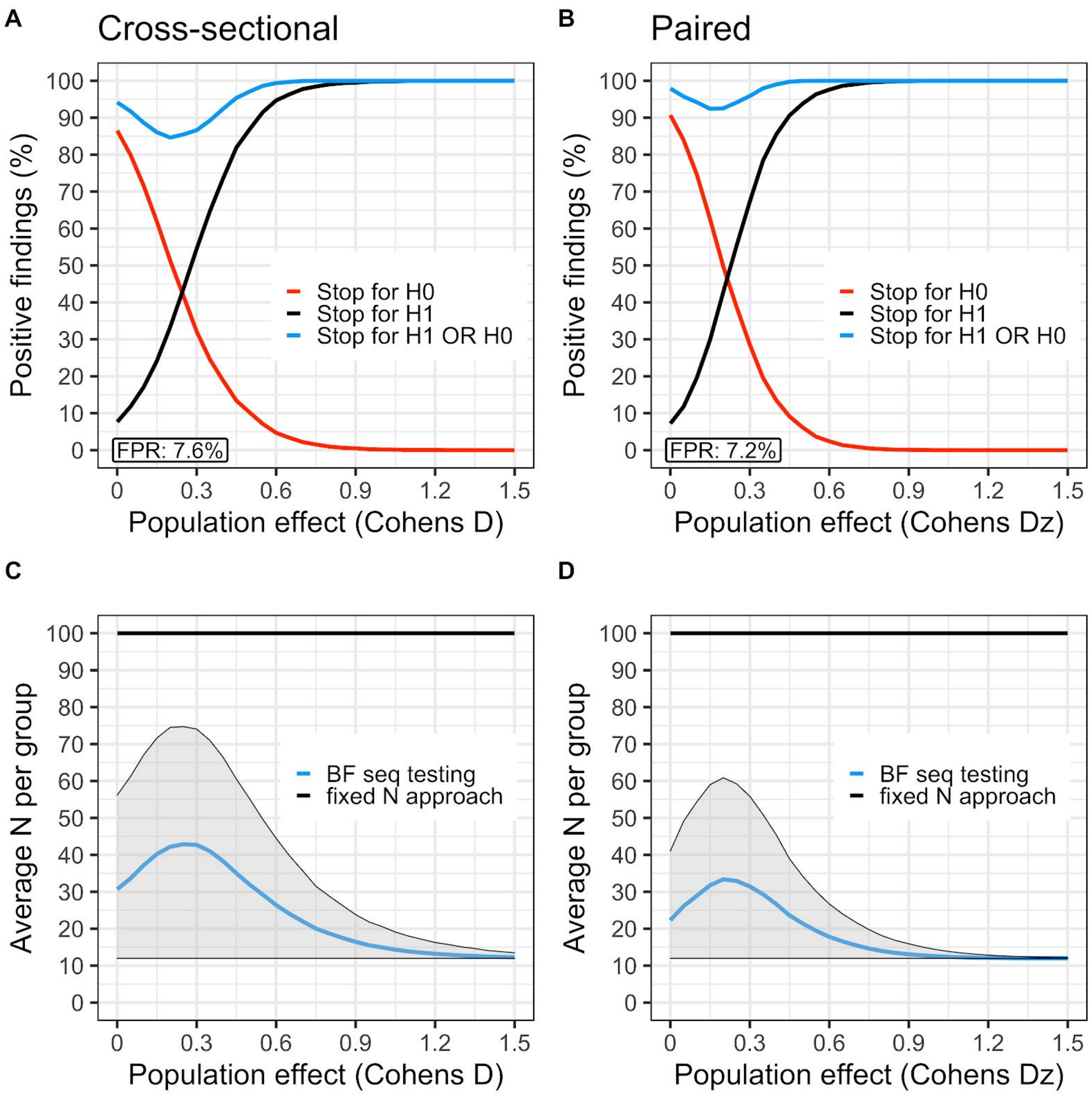


*Figure S9. Settings for the simulation: H1 is a one sided cauchy(0,0.707), (Nstart = 12, Nmax = 100, BF threshold = 4). A and B) The black curve shows the proportion of studies that showed support for H1 (BF>4) during data collection, at a range of population effects (starting at no effect, D = 0). The red curve is the proportion of studies showing support for H0 (BF<¼). The blue curve is the sum of the red and black curves. C and D) shows the average number of subjects needed to reach a stopping decision at different population effects. The flat black line represents Nmax (100 subjects/group). BF sequential testing is the blue line with shaded area denoting ± 1 SD.*

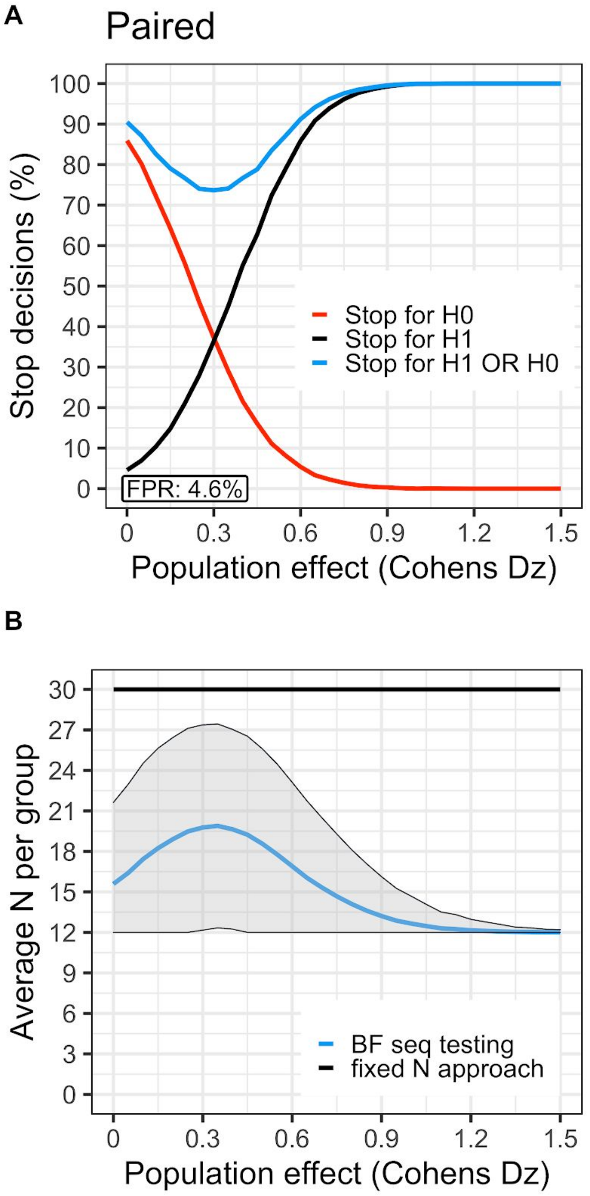


*Figure S10. Settings for the simulation: H1 is a one sided cauchy(0,1), (Nstart = 8, Nmax = 30, BF threshold = 4). A ) The black curve shows the proportion of studies that showed support for H1 (BF>4) during data collection, at a range of population effects (starting at no effect, D = 0). The red curve is the proportion of studies showing support for H0 (BF<¼). The blue curve is the sum of the red and black curves. B) shows the average number of subjects needed to reach a stopping decision at different population effects. The flat black line represents Nmax (30 subjects/group). BF sequential testing is the blue line with shaded area denoting ± 1 SD. The fact that a lower average sample size trades off against a higher risk of false negatives can be seen by comparing this figure to panel B and D in Figure 5 from the main text which have the same settings except for: 1)* $H1 : \delta \sim Cauchy(0, 0.707)$ *instead of* $H1 : \delta \sim Cauchy(0, 1)$ *and 2) testing starts at 12 subjects.*

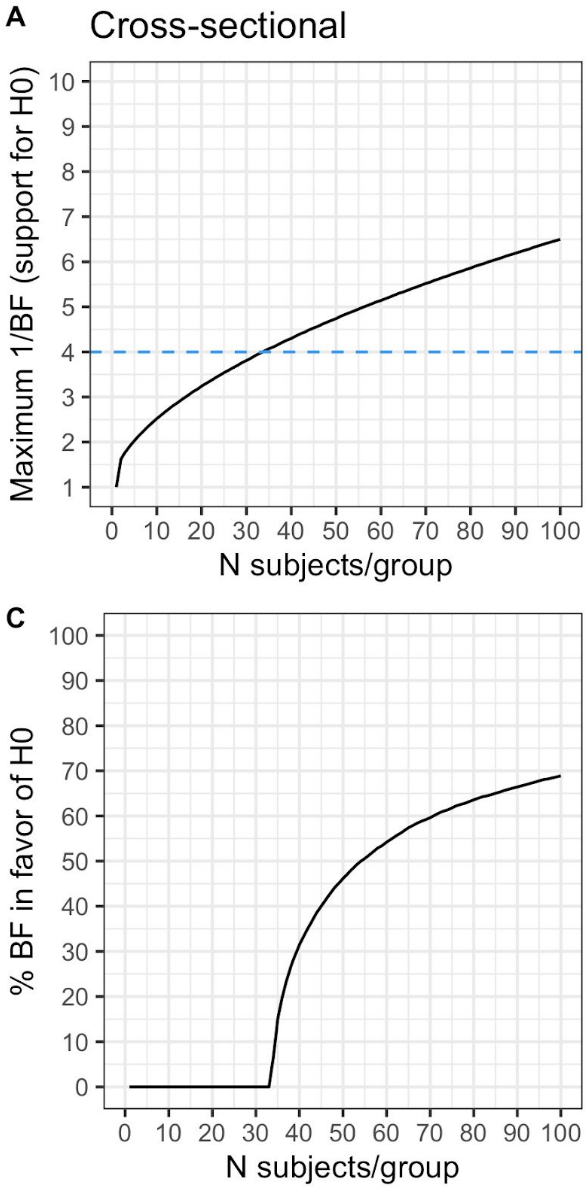


*Figure S11. Maximum possible support (1/BF) in favor of H0 compared to H1 when using a two-tailed BF t-test. A-D) When using the settings described in the main article (H1 is specified as a Cauchy(0,0.707), Nstart = 12, threshold = 4) but a two-tailed test instead of a one-tailed test, it is not possible to obtained evidence in favor of H0 at smaller N. E.g., in a cross-sectional design, at least 34 subjects/group are needed before the BF can reach a threshold of ¼. C-D) Percentage of BF showing support (1/BF > 4) for H0 at different N. E.g., at 50 subjects/group, only 45% of BF will show support in favor of H0, when H0 is true. Hence, in order to stop for H0 when using commonly seen sample sizes in PET studies, we recommend to use a one-tailed BF t-test instead. This means that the researchers must make a prediction of the direction of the effect before initiating the study.*

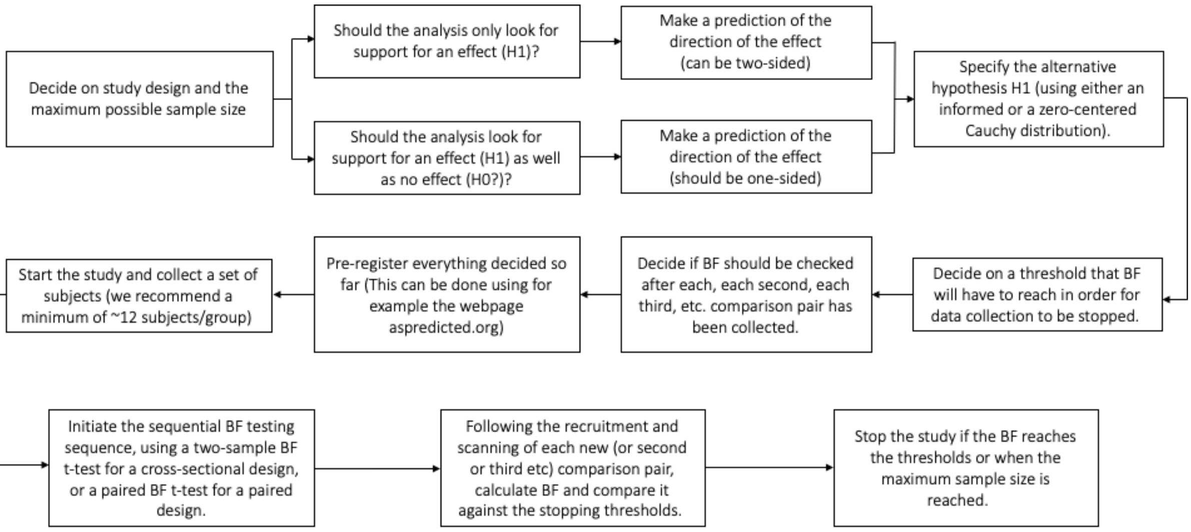


*Figure S12. Recommended steps to follow in order to perform a clinical PET study using sequential BF testing, for a paired or cross-sectional design.*
